# LifeSim: A Lifecourse Dynamic Microsimulation Model of the Millennium Birth Cohort in England

**DOI:** 10.1101/2021.02.12.21251642

**Authors:** Ieva Skarda, Miqdad Asaria, Richard Cookson

## Abstract

We present a dynamic microsimulation model for childhood policy analysis that models developmental, economic, social and health outcomes from birth to death for each child in the Millennium Birth Cohort (MCS) in England, together with public costs and a summary wellbeing measure. The model is a discrete event simulation in discrete time (annual periods), implemented in R, which progresses 100,000 individuals through each year of their lives from birth in the year 2000 to death. From age 0 to 18 the model draws observational data from the MCS, with explicit modelling of only a few derived outcomes (mental health, conduct disorder, mortality, health-related quality of life, public costs and a general wellbeing metric). During adulthood, all outcomes are modelled dynamically using explicit networks of stochastic process equations, with separate networks for working age and retirement. Our equations are parameterised using effect estimates from existing studies combined with target outcome levels from up-to-date administrative and survey data. We present our baseline projections and a simple validation check against external data from the British Cohort Study 1970 and Understanding Society survey.

## 1 Introduction

Recent scientific advances have established beyond reasonable doubt that childhood programmes can have important effects on health and wellbeing many decades in the future, during working years and retirement (Conti, Mason and Poupakis, 2019; Heckman, 2012). Policy makers want quantitative information about these long-term effects, and they also want information about distributional impacts on inequality in lifetime health and wellbeing, as well as inequality in current period health and wellbeing. While childhood policy analysis using randomised control trials and quasi-experiments is the gold standard in establishing cause and effect relationships, this is rarely possible when quantifying lifetime policy effects over many decades. Even when long-term follow-up data is available, such analysis yields insights about historical cohorts born many decades ago with questionable relevance to the current childhood policy context. Microsimulation offers a forward-looking alternative for childhood policy analysis, as it can extrapolate long-term outcomes for cohorts living in the present and project the effects of the policies that policymakers are considering today.

In this paper we introduce a dynamic childhood policy microsimulation model “LifeSim” which models the co-evolution of economic, social and health outcomes from birth to death for each child in a general population birth cohort of 100,000 English children born in year 2000-1. In addition to modelling the individual outcomes, LifeSim also models the associated costs and savings to the public budget.

The chosen life outcomes and the structure of our model are designed to address cross-sectoral childhood policy concerns and to align with the large body of theory and knowledge about human capital formation in childhood and later life economic and health outcomes. From age 0 to 18 we heavily rely on observed life outcomes from the Millennium Cohort Study (MCS), and only explicitly model three specific childhood outcomes - mental health, conduct disorder and mortality - which are then combined with MCS data to estimate public costs and a general wellbeing metric. During adulthood, however, we specify explicit networks of stochastic processes, with different networks for working years and retirement, and parameterise these using estimates from published studies of longitudinal data on earlier cohorts.

LifeSim has the following distinct features:

i. it jointly models the co-evolution of many economic, social and health outcomes, capturing how outcomes in multiple domains interact, compound and cluster over time, emphasising how early-life disadvantages can compound over life creating a spiral of multiple disadvantage;
ii. it simulates long-run outcomes for a whole general population cohort of children, not just one specific subpopulation of trial participants, which allows the model to serve as a platform for many different kinds of informative policy analysis, including optimal policy targeting analysis, population-wide distributional impact analysis and assessment of the opportunity costs falling on the individuals not directly affected by the intervention;
iii. it simulates individual-level outcomes for each heterogeneous child in the cohort, instead of only producing average-level outcomes, allowing us to produce multidimensional individual wellbeing measures, which have been discussed in the literature and have well-known advantages over un-weighted cost-benefit analysis (Adler and Fleurbaey, 2016);
iv. it simulates outcomes over the whole lifecourse from birth to death, enabling policy analysis to adopt a broad lifetime perspective.

We capture all of these features by combining many different sources of data, which requires strong assumptions. We make all of our assumptions explicit and subject to scrutiny by providing carefully labelled and fully referenced details of all modelling equations, parameters and data sources in the appendix, and by publishing our complete programming code. We use longitudinal data on children born in 2000 as our primary data source but supplement this with other sources of data including more up-to-date cross-sectional administrative and survey data as well as older sources of longitudinal data on children born in earlier decades. In choosing how many assumptions to make and how many sources of data to use, there are trade-offs between internal and external validity.^1^ Using a single source of experimental data with long-term follow-up over many decades would maximise internal validity, but is only possible for backward-looking evaluation of policy experiments many decades ago. Using assumptions and multiple sources of data is necessary to achieve external validity for forward-looking economic appraisal of current policy options in the current policy environment.

To our knowledge LifeSim is the first microsimulation model that provides information on many developmental, economic, social, health and public cost outcomes from birth to death for each individual in a birth cohort. In the economics literature, there are dynamic microsimulation models of many co-evolving economic and social outcomes across the life-cycle (e.g. LINDA, a rational agent dynamic microsimulation based on dynamic programming, Van der Ven (2016)) and dynamic microsimulation models of childhood development (e.g. MELC, a discrete event simulation from age 0 to 13, Milne et al. (2015)). And in the health literature there are dynamic microsimulation models of multiple co-evolving health and public cost outcomes (e.g. HealthPaths, Wolfson and Rowe (2014), POHEM, Hennessy et al. (2015) and IMPACT NCD Kypridemos et al. (2016)). However, none of these cover developmental, economic, social and health outcome domains and few provide information on the whole lifecourse from birth to death.^2^ Modelling the entire lifecourse allows us to examine how childhood outcomes can lead to spirals of advantage and disadvantage in later life, whereby economic, social and health outcomes interact, compound and cluster over time. For example, a young child with poor cognitive and social skills is at heightened risk of multiple adverse outcomes as they grow older – including unhealthy behaviour, mental illness, unemployment, low earnings imprisonment and physical illness – all of which can interact and compound in a spiral of disadvantage (Zucchelli, Jones and Rice, 2012; Layard et al., 2014; Frijters, Bellet and Krekel, 2017). Modelling this also provides a platform for more informative long-term economic evaluation, targeting analysis and distributional analysis of childhood policies from a lifetime perspective, as we illustrate in a companion paper under review elsewhere (Skarda, Asaria and Cookson, 2021).

## 2 Methods

### 2.1 Model Structure

Our microsimulation model is a discrete event simulation in discrete time (annual periods), which progresses 100,000 individuals through each year of their lives from birth in the year 2000 to death. From ages 0 to 18 it closely follows observed Millennium Cohort Study (MCS) data, and thereafter predicts the annual evolution of each life outcome based on the current values of relevant characteristics and outcomes, which in term depend on lagged values.^3^ This kind of model can be seen as a pragmatic compromise between a simpler Markov model structure, which has no “memory” or dependence upon lagged values, and a more complicated agent-based model structure, which explicitly models interactions between individuals and how individual behaviour may depend upon the macro-level policy environment as well as the behaviour of others. Allowing dependence upon lagged values allows a rich analysis of the dynamic clustering and compounding of multiple outcomes over time, while setting aside agent-based interactions keeps the model tractable, even when modelling a relatively large number of outcomes.

The model links together a diverse set of individual-level life outcomes of interest to policymakers (Figure 1). By using rich observational data from the MCS, our model provides information on various aspects of human capital development in childhood - including social skills, cognitive skills and health behaviour (teenage smoking) - and then extrapolates later life outcomes across economic, social and health domains for the rest of the lifecourse. For simplicity and concreteness we focus on one important and readily measurable dimension of social skills - conduct problems - as proxied by two separate parent reported measures. Child conduct is related to self-control and regulation, which have been shown to matter in many aspects of life, including wellbeing, income, employment, crime and health outcomes (Goodman et al., 2015). We also model mental illness and health-related quality of life during childhood, using external datasets (Mental Health of Children and Young People Great Britain, and a dataset by Love-Koh et al. (2015)).

**Figure 1:**
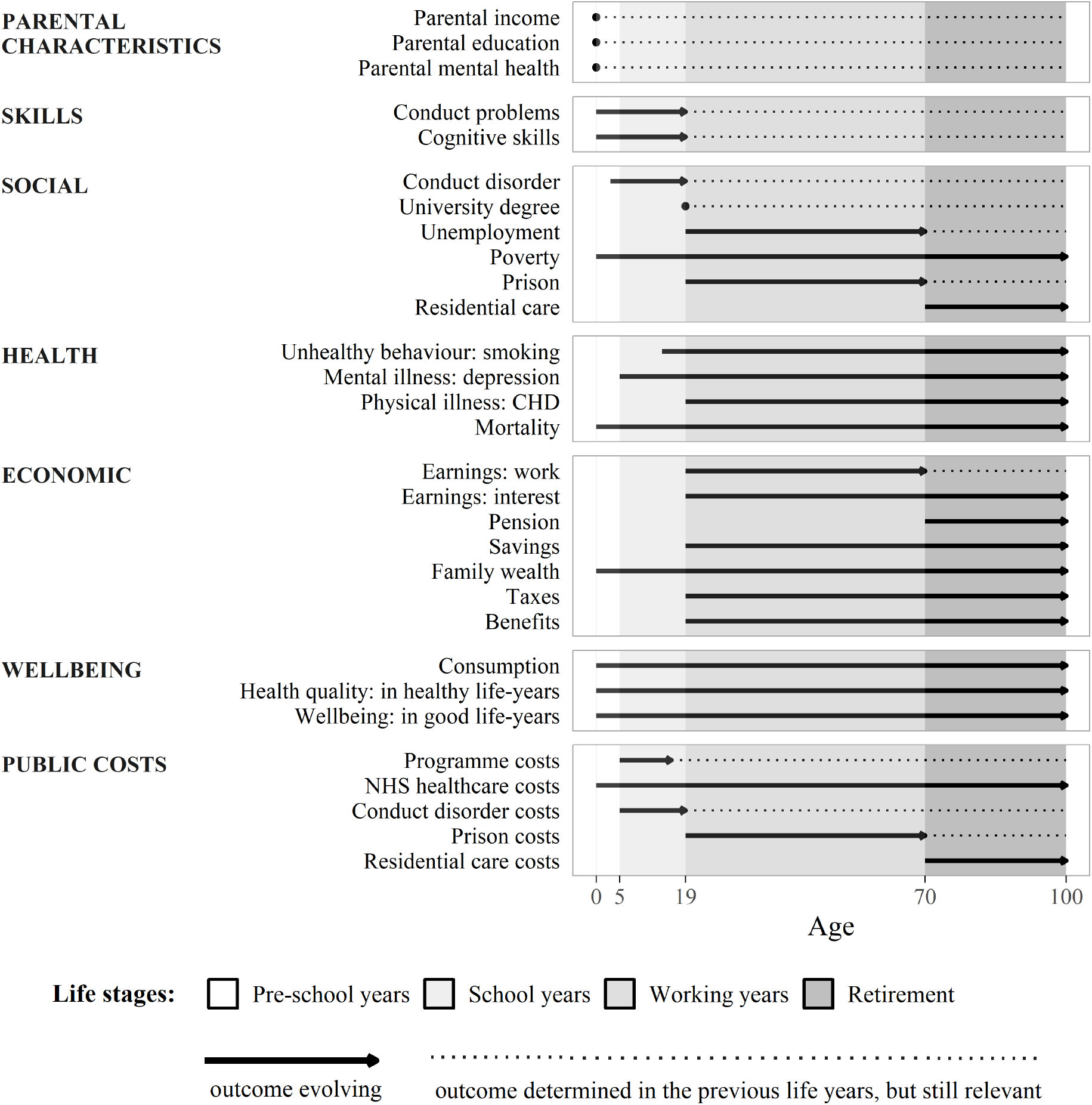
Overview of Key Outcomes Over the Lifecourse.

Let *i* = 1, ..100, 000 index the individual children in the cohort. Let yearly time periods also corresponding to the age of children be indexed as *age* = 0, 1, ..18..*T* where *age* = 18 marks the end of childhood, and *T* is the last time period in which there are any cohort members still alive (which we assume to be 100, since small number problems make predictions decreasingly reliable at older ages). Let *X*_*i*_ be the vector of initial conditions assumed to be constant for child *i* (e.g. individual and family characteristics at birth or other early time period – if data at birth on the condition is not available); let *θ*_*i,age*_ be an age-specific vector of child Strengths and Difficulties Questionnaire (SDQ) scores – multi-dimensional parent-reported score on child’s difficulties, *cd*_*i,age*_ – an age-specific outcome of whether child develops a conduct disorder, and *cog*_*i,age*_ – an age-specific child’s cognitive skills measure. Finally, let *Y*_*i,age*_ be the age-specific vector of life-cycle outcomes (further, outcomes) for child *i*. These outcomes can be further classified as social, health and economic outcomes, i.e. *Y*_*i,age*_ ≡ {*S*_*i,age*_, *H*_*i,age*_, *E*_*i,age*_}, where *S*_*i,age*_, *H*_*i,age*_, *E*_*i,age*_ are the vectors of social, health and economic outcomes respectively. It is allowed for the vector *X*_*i*_ to also contain elements of {*S*_*i*_, *H*_*i*_, *E*_*i*_}.

At each age the individual probability of dying *pr*.*dead*_*i,age*_ is modelled and defined over the closed interval from zero to one, i.e. *pr*.*dead*_*i,age*_(*E*_*i,age*_, *S*_*i,age*_, *H*_*i,age*_) ∈ [0, 1], which then determines the discrete outcome *dead*_*i,age*_ – whether the individual at a certain age is dead (*dead*_*i,age*_ = 1) or alive (*dead*_*i,age*_ = 0). More specifically, we can represent the outcome ‘dead or alive’ by a function *l*(.) such that if in the previous year individual was alive then they can be either alive or dead in the following year, i.e. 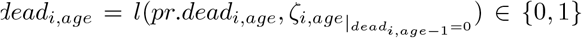 (where *ζ*_*i,age*_ represents stochasticity); and if in the previous year individual was dead then, because death is an ‘absorbing state’, they can be only dead in the following year, i.e. 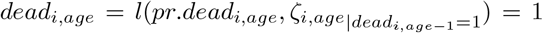. Individual life span is then 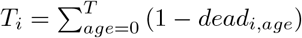.

To describe the initial conditions (in vector *X*_*i*_), we draw observations on each child 100,000 times in total to represent a cohort of 100,000 individuals, using re-sampling with replacement from the initial sweeps of MCS – a longitudinal survey of English children born in 2000-2001 (see panels A-C of table 1).

**Table 1:** Summary of Child Characteristics and Family Conditions.

| Initial condition | Data used in modelling | Source | Mean | SD | Min | Max |
| --- | --- | --- | --- | --- | --- | --- |
| <u>A: Basic child's characteristics</u> |  |  |  |  |  |  |
| Sex | Indicator if child is male | MCS1 | 0.49 | 0.50 | 0 | 1 |
| Teenage smoking | Indicator if child smokes at 14 | MCS6 | 0.16 | 0.37 | 0 | 1 |
| <u>B: Social conditions</u> |  |  |  |  |  |  |
| Parental income | OECD equivalised household income after taxes and benefits, £ | MCS1 | 32,004 | 19,972 | 1,445 | 128,246 |
| Parental wealth | Parental assets, £ | MCS5 | 3,068 | 19,937 | 0 | 600,000 |
| Parental socio-economic position | Household income quintile | MCS1 | 3.06 | 1.37 | 1 | 5 |
| Childhood poverty | Indicator if household income is below 60% median | MCS1 | 0.27 | 0.45 | 0 | 1 |
| <u>C: Parental characteristics</u> |  |  |  |  |  |  |
| Parental education | Indicator if parent has a university degree (NVQ 4 or above) | MCS1 | 0.31 | 0.46 | 0 | 1 |
| Parental depression at child's birth | Indicator if Rutter malaise inventory score is 4 or above | MCS1 | 0.14 | 0.35 | 0 | 1 |
| Parental depression severity at child's birth | 9-item Rutter malaise inventory score | MCS1 | 1.66 | 1.68 | 0 | 9 |
| Parental depression when child is 5 years old | Indicator if Kessler psychological distress scale score is 13 or above | MCS3 | 0.03 | 0.18 | 0 | 1 |
| Parental depression severity when child is 5 years old | 6-item Kessler psychological distress scale score | MCS3 | 3.17 | 3.72 | 0 | 24 |
| <u>D: Child's social skills</u> |  |  |  |  |  |  |
| Conduct problems up to age 4 | SDQ conduct problem score (see the note) | MCS2 | 2.87 | 2.02 | 0 | 10 |
| Conduct problems, ages 5-6 | SDQ conduct problem score | MCS3 | 1.46 | 1.47 | 0 | 8 |
| Conduct problems, ages 7-10 | SDQ conduct problem score | MCS4 | 1.32 | 1.50 | 0 | 9 |
| Conduct problems, ages 11-13 | SDQ conduct problem score | MCS5 | 1.38 | 1.58 | 0 | 10 |
| Conduct problems, age 14+ | SDQ conduct problem score | MCS6 | 1.42 | 1.68 | 0 | 10 |
| Impact of problems up to age 4 | SDQ impact supplement score | MCS2 | 0.11 | 0.58 | 0 | 8 |
| Impact of problems, ages 5-6 | SDQ impact supplement score | MCS3 | 0.13 | 0.63 | 0 | 8 |
| Impact of problems, age 7+ | SDQ impact supplement score | MCS4 | 0.25 | 0.91 | 0 | 9 |
| <u>E: Child's cognitive skills</u> |  |  |  |  |  |  |
| Cognitive skills up to age 4 | Various measures (see the note) | MCS2 | 1.02 | 0.14 | 0.58 | 1.44 |
| Cognitive skills, ages 5-6 | Various measures | MCS3 | 1.03 | 0.13 | 0.57 | 1.45 |
| Cognitive skills, ages 7-10 | Various measures | MCS4 | 1.03 | 0.14 | 0.56 | 1.41 |
| Cognitive skills, ages 11-13 | Various measures | MCS5 | 1.03 | 0.15 | 0.39 | 1.49 |
| Cognitive skills, age 14+ | Various measures | MCS6 | 1.02 | 0.14 | 0.43 | 1.50 |

Note: The analysis is for 100,000 individuals in the LifeSim cohort. MCS $j$ denotes MCS sweep $j$ (6 sweeps in total). Children were 9 months old in MCS1, 3 years old in MCS2, 5 years old in MCS3, 7 years old in MCS4, 11 years old in MCS5 and 14 years old in MCS6. SDQ conduct problem and impacts scores have a scale 0-10, with a higher value representing more problems/higher impact of problems. The cognitive skills measure is an age-specific common factor extracted from the various cognitive skills measures in MCS, including the British Ability Scales II, Bracken School Readiness Assessment, National Foundation for Educational Research Progress in Maths, Cambridge Neuropsychological Test Automated Battery tests and Applied Psychology Unit.

Similarly, we use MCS data from all of the sweeps (up to age 14) to collect data for the vector *θ*_*i,age*_ – information on child SDQ conduct problem subscale score and a further parent-reported “behavioural impact” score (see panel D of table 1). Both of these scores range from 0-10, with a higher score representing more conduct problems and a more severe impact of difficulties in child’s life. MCS data are reported at sweeps every 2 to 4 years, so we use the most recent MCS sweep data available to fill in the missing values in the time gaps, and for age 15-18.

We then use the reported SDQ score components to model whether or not a child develops conduct disorder, using a previously developed algorithm which predicts a child’s probability of developing conduct disorder as a function of SDQ score components: the SDQ conduct problem score and the “behavioural impact” score. This provides a specific probability of conduct disorder based on a classification as either “possible” or “probable” (Goodman et al., 2003; Goodman, Renfrew and Mullick, 2000).^4^ This modelled probability is then combined with a random draw from a uniform distribution over 0-1, which allows us to simulate the discrete outcome of whether or not a child develops conduct disorder. Formally, the age-specific conduct disorder outcome *cd*_*i,age*_ can be represented as:

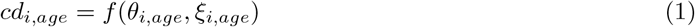

where *ξ*_*i,age*_ represents stochasticity.

We also use the MCS data from later sweeps (up to age 14) to build *cog*_*i,age*_ – a single measure of each child’s cognitive skills at each age throughout their childhood up to age 18 (see panel E of table 1). More specifically, our cognitive skills measure is an age-specific common factor extracted from the cognitive skills measures available in MCS, including the British Ability Scales II (for ages 3, 5, 7, 11), Bracken School Readiness Assessment (for age 3), National Foundation for Educational Research Progress in Maths (for age 7), Cambridge Neuropsychological Test Automated Battery tests (for ages 11 and 14) and Applied Psychology Unit (for age 14). We extract a common factor for each age where test results are available using principal component analysis, and standardise it to be with a mean of 1.00 and standard deviation of 0.15 (following Jones and Schoon (2008)). Similar to the SDQ score data, we use the most recent MCS sweep data available to fill in the missing values in the time gaps, and for age 15-18.

During adulthood, child’s SDQ scores, conduct disorder outcomes and cognitive skills are assumed to stay fixed at the level achieved by the end of childhood, i.e. *θ*_*i,age*_ = *θ*_*i*,18_, *cd*_*i,age*_ = *cd*_*i*,18_ and *cog*_*i,age*_ = *cog*_*i*,18_ for *age* = 19..*T*_*i*_.

Over the life-cycle (*age* = 0..18..*T*_*i*_), the vector of other life-cycle outcomes *Y*_*i,age*_ evolves as:

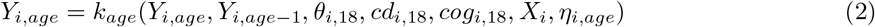

where *η*_*i,age*_ represents stochasticity. It should be noted that separate outcomes in the vector *Y*_*i,age*_ can depend on a subset (and not necessarily all) of the outcomes in the vectors *Y*_*i,age-*1_, *Y*_*i,age*_, *θ*_*i*,18_, *cd*_*i*,18_, *cog*_*i*,18_, *X*_*i*_, which can be achieved by restricting coefficients. Also, a period-specific outcome in the vector *Y*_*i,age*_ will generally not depend on itself, but can depend on other outcomes at that time period included in the vector *Y*_*i,age*_.

The model structure specified by *k*_*age*_(.) changes as individuals progress through key life stages. In each life stage, the dependencies between the initial conditions and the life-course outcomes are represented by model structure diagrams in Figure 2 and Figure 3, and are also summarised in Table 2. In the model structure diagram each solid arrow is modelled using equations (as we will explain in more detail in Section 2.3).

**Table 2:**
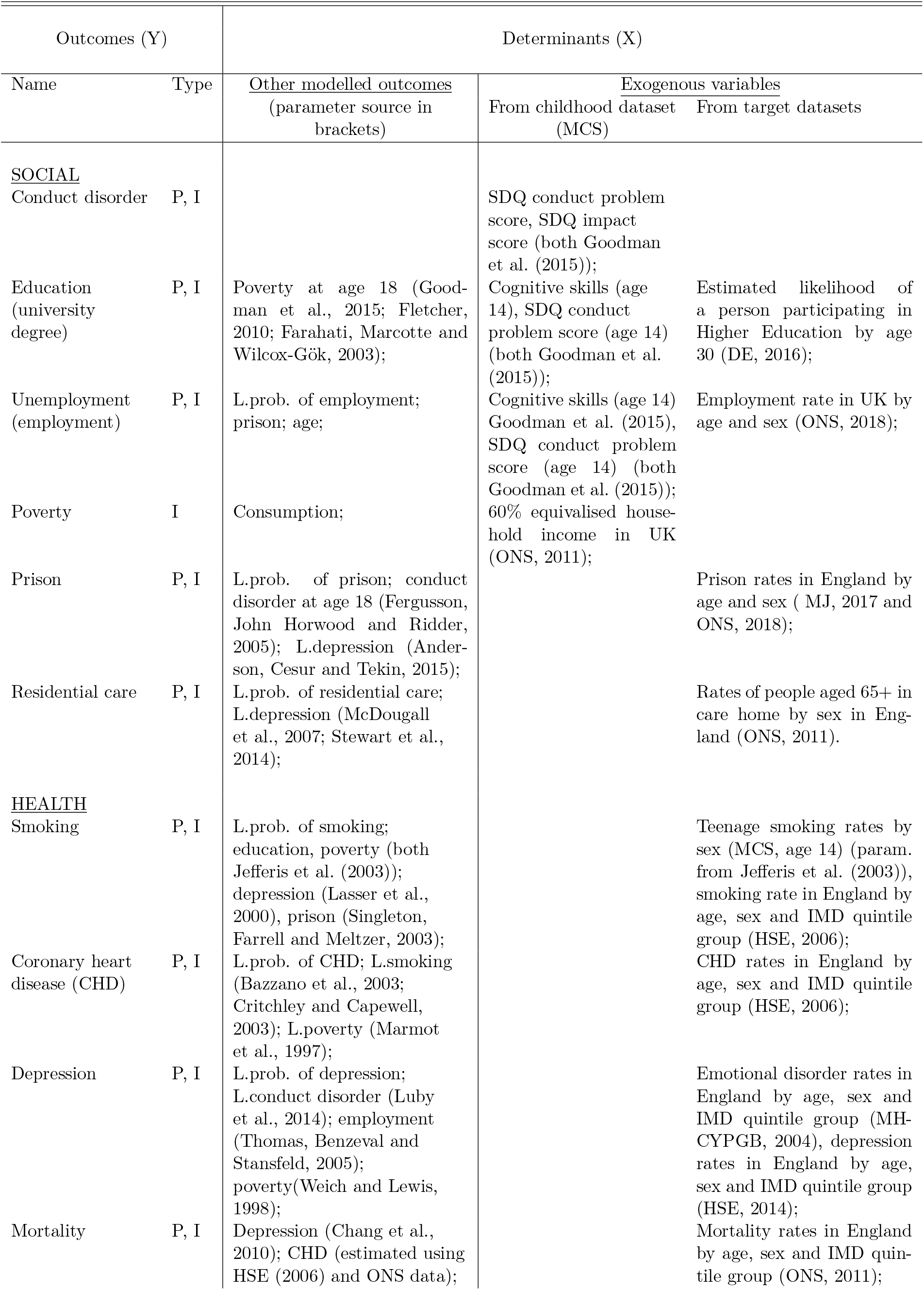
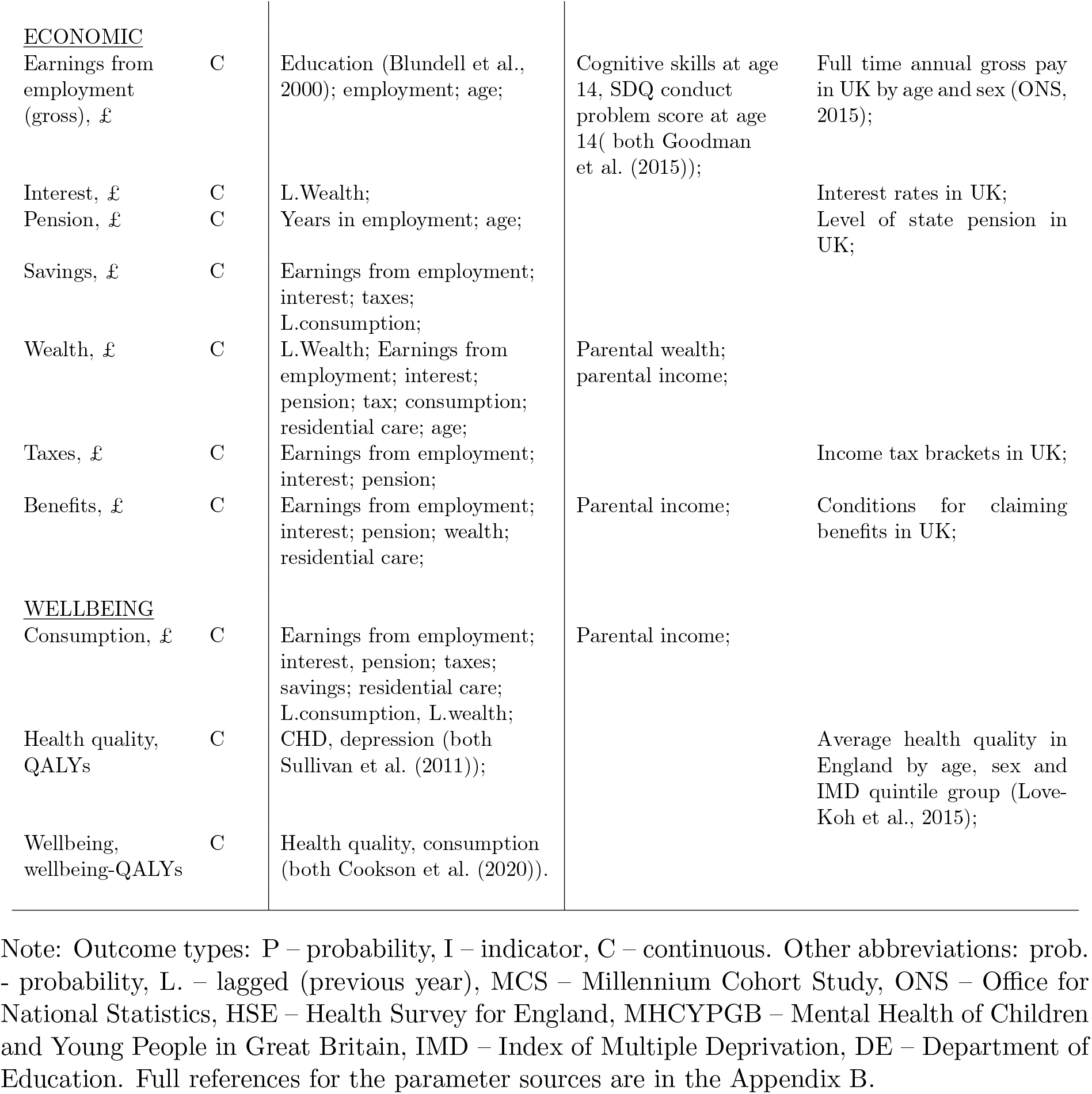
Determinants of the Modelled Outcomes.

**Figure 2:**
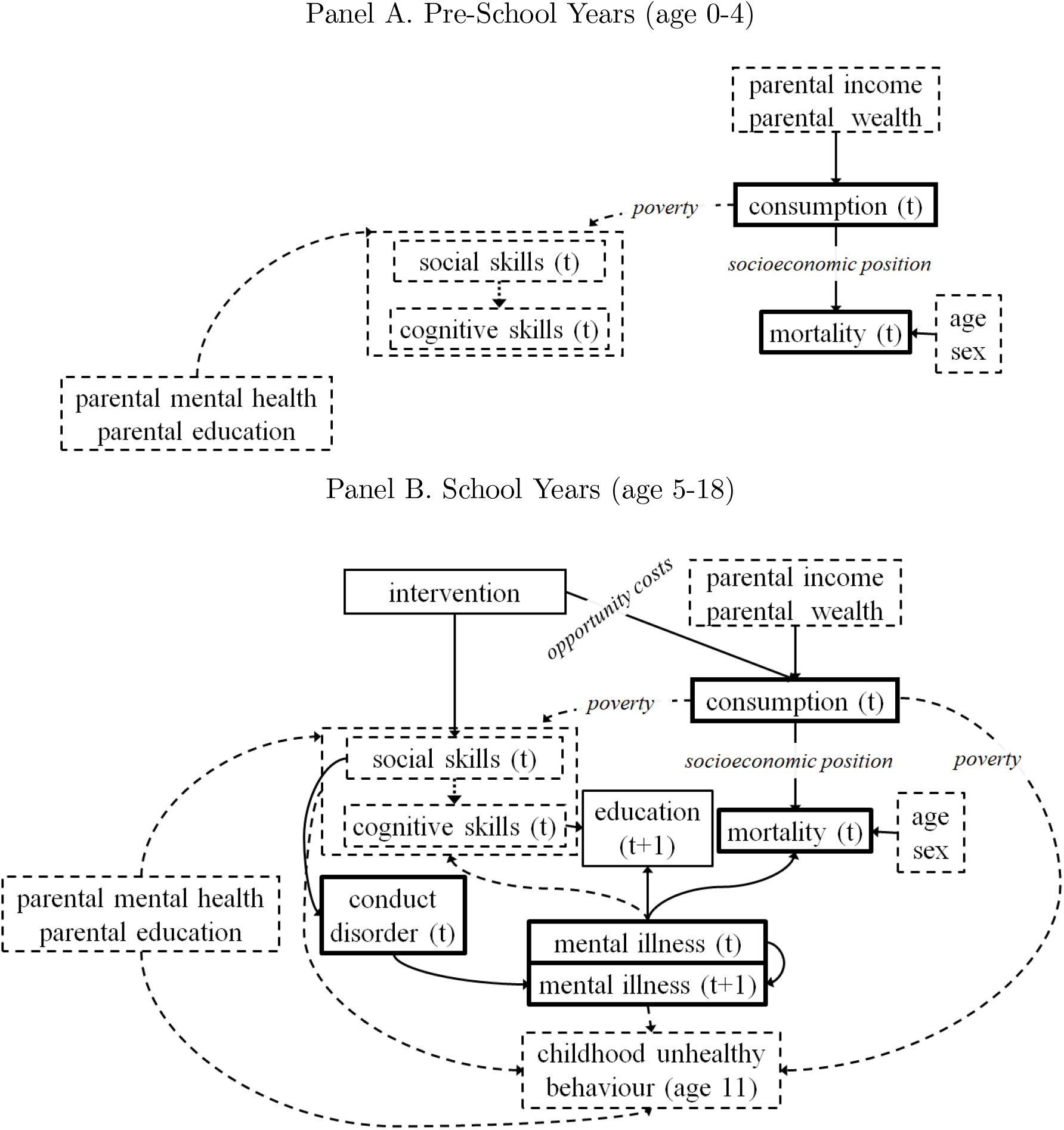
Model Structure for Key Life Stages in Childhood. Note: The solid arrows represent the stochastic processes that we model using modelling equations as described in Section 2.3; the dashed arrows represent implicit processes that we do not explicitly model, but that exist in the childhood dataset. The boxes represent life outcomes: the dashed boxes represent exogenous inputs into our model that are taken as given either from the childhood dataset or the previous life-stage, the thick boxes represent final outcomes that directly influence wellbeing or impose a cost to the public budget.

**Figure 3:**
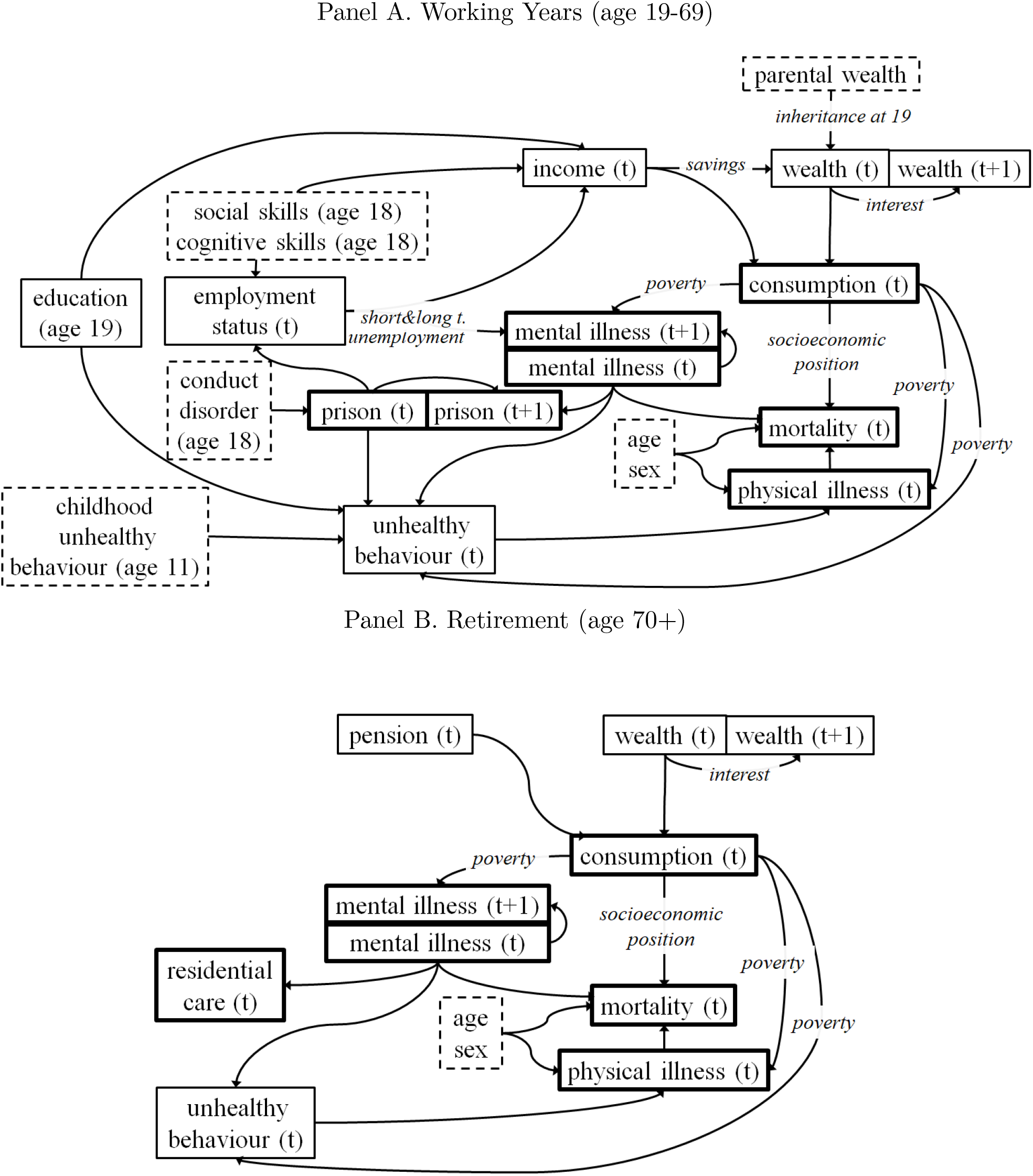
Model Structure for Key Life Stages in Adulthood. Note: The solid arrows represent the stochastic processes that we model using modelling equations as described in Section 2.3. The boxes represent life outcomes: the dashed boxes represent exogenous inputs into our model that are taken as given either from the childhood dataset or the previous life-stage, the thick boxes represent final outcomes that directly influence wellbeing or impose a cost to the public budget.

In choosing the model outcomes and formulating the model structure we consulted with experts in childhood development and childhood policy, demography, epidemiology, human capital economics and labour economics (see list of advisory group members in the acknowledgements) and were also guided by inter-disciplinary theory on human capital formation in childhood and how this influences educational attainment, earnings, physical illness, mental illness, mortality and other outcomes with important impacts on individual wellbeing and public cost (Almond, Currie and Duque, 2018; Goodman et al., 2015; Nelson et al., 2020; Cunha and Heckman, 2010; Adler and Stewart, 2010; O’Donnell, Van Doorslaer and Van Ourti, 2015; Layard et al., 2014; Shonkoff, 2010; Black et al., 2017).

LifeSim also models variables relevant to the public budget (Figure 4). This includes modelling the public costs over time associated with certain life outcomes, such as conduct disorder, being in prison, mental illness, coronary heart disease, as well as cash benefits paid to people who are in poverty and/or unemployed. This also includes modelling the taxes paid over time on individual earnings and financial gains. These can be aggregated, to assess the overall impact on the public budget as well as cost savings under different policy scenarios and over various time spans. Details of the evidence and assumptions about the unit costs of public services and our simple approach to modelling long-run taxes and benefits are found in Appendix A.

**Figure 4:**
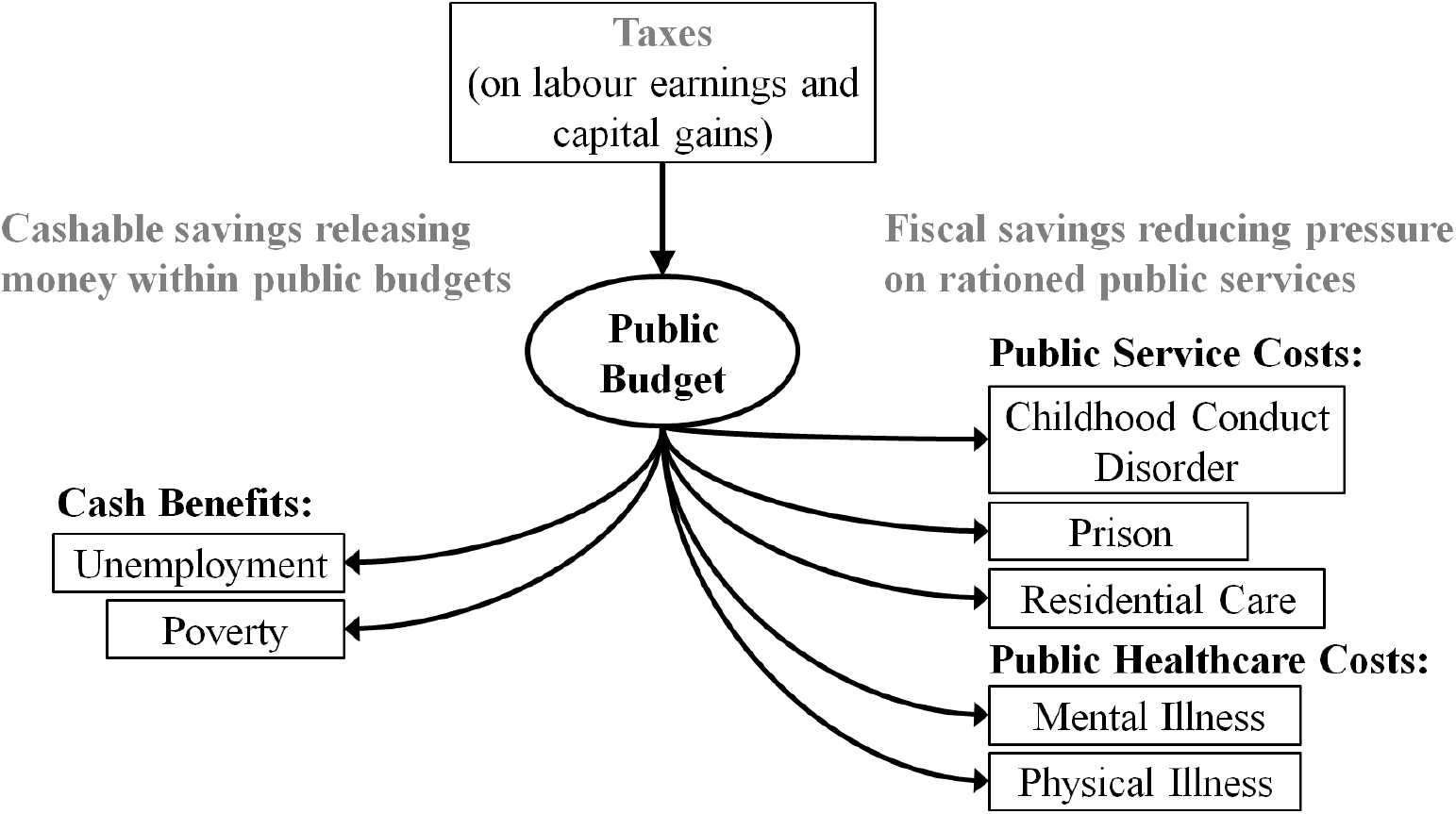
Model Structure for Public Costs.

### 2.2 Parameters

To model later life outcomes, we use equations parameterised using (i) cross-sectional target data which describe expected levels of and associations between variables at a point in time, usually based on up-to-date survey or administrative data, and (ii) effect estimates which attempt to draw inferences about the effect of one variable on another variable, either at the same time or a future point in time, usually based on statistical analysis of longitudinal data on historical cohorts. Our target data comes from recent and nationally representative available surveys and administrative records in England. Our effect estimates come from studies based on longitudinal data in a UK context, unless robust estimates are only available from other high-income countries. Our effect estimates come from studies of longitudinal data which control for observed confounding factors and focus on plausible causal relationships for which there is a large body of theoretical and empirical evidence. Nevertheless, our estimates are subject to potential omitted variable bias and cohort bias. For example, we take the estimated effect of childhood SDQ score on earnings in young adulthood from a study of longitudinal data on children born in 1970, which controls for observed child-level, family-level and neighbourhood-level factors. We interpret this as a causal estimate i.e. if you increase SDQ score you will increase adult earnings by this amount. However, this estimate may be too low or too high if there are unobserved variables which influence both SDQ score and earnings (“omitted variable bias”). It may also be biased if the underlying stochastic processes have changed since 1970, such that SDQ score is now a more or less powerful determinant of adult earnings (“cohort bias”). Using estimates based on past cohorts of individuals thus relies on the assumption that micro-level causal effects do not change much over many decades (e.g. the proportional effect of social skills on earnings for an individual), even though the macro-level prevalence of each outcome within society may change dramatically (e.g. the average levels of social skills and earnings).

Table 2 summarises the dependencies between the modelled outcomes together with parameter sources for effects estimates, if applicable, as well as the dependencies of the modelled outcomes on the target datasets and the variables from the MCS childhood dataset. More details, as well as the full description of the target datasets are found in Appendix A.

### 2.3 Modelling Equations

Most of the equations modelling the outcomes can be described as one of the following: (i) simple level equations based on target data only; (ii) complex level equations based on target data supplemented with effect estimates; (iii) simple difference equations based on age associations observed in cross-sectional target data; (iv) complex difference equations based on age associations observed in cross-sectional target data supplemented with effect estimates. We illustrate each below in turn with a simple example. We also use equations that do not fit this taxonomy to model specific variables, such as savings behaviour and wealth accumulation over time, as well as public costs (more on this can be found in Appendix A).

#### Level Equations

To model the individual probability of dying, the simplest approach is to use historical mortality rates:^5^

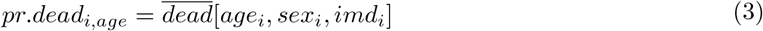

where 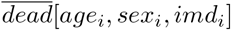 is the mean probability of dying conditional on age, sex and English index of multiple deprivation (IMD) quintile group, calculated using a target dataset such as the Office for National Statistics mortality data (see Table A.2). We denote means from a target dataset using an overline.

We can also supplement equation (3) with effects estimates. For example, we may wish to model that coronary heart disease (CHD) increases one’s probability of dying by a certain proportion (denoted by 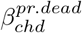). In this case, we use:

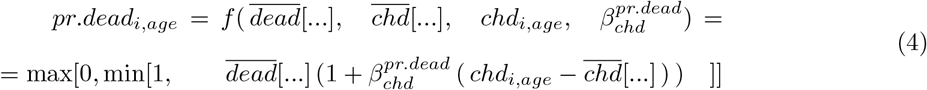

where *chd*_*i,age*_ is the simulated binary outcome of individual *i* having a CHD at a certain age, 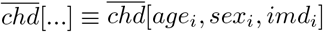 is the mean CHD prevalence given age, sex and IMD quintile group from a target dataset. Notice that we subtract the mean CHD prevalence from the simulated CHD outcome to avoid double counting, as the term 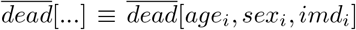 is not independent from CHD, but the variable CHD is not observable in the ONS mortality target dataset, so we cannot directly condition the target mortality mean on the CHD status. After multiplying each term in the brackets by the beta coefficient, it can be seen that our approach is equivalent to subtracting the ‘population attributable risk’ from the risk of the simulated individual (Webb, Bain and Page, 2016).

#### Difference Equations

If a level of a variable is already known, we can proceed by modelling the evolution of a variable as a difference from a previous time period. For example, when the level of earnings has been established at age 19 (the start of ‘working years’ life stage), we can model the change in individual earnings during the subsequent periods as:

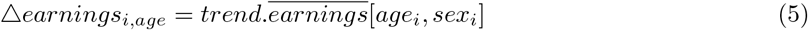

where Δ*earnings*_*i,age*_ = *earnings*_*i,age*_ - *earnings*_*i,age-*1_ is the change in earnings from the previous year, and 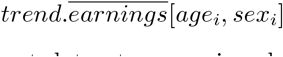 is a trend that governs the changes in earnings over time, calculated from a target dataset on earnings by age and sex.

Similar to level-equations, we can supplement equation (6) with an effect estimate. For example, to model that developing depression reduces earnings by a certain level represented by 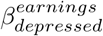 we use:

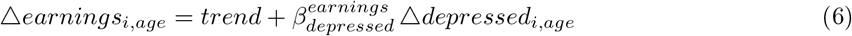

where *depressed*_*i,age*_ is an indicator of an individual having a depression at a given age and Δ*depressed*_*i,age*_ = *depressed*_*i,age*_ - *depressed*_*i,age-*1_ More details on the modelling equations are found in Appendix A.

### 2.4 Wellbeing Summary Measure

Conventional methods of unweighted benefit-cost analysis do not provide direct information about impacts on wellbeing and can be criticised on two important grounds. First, by focusing on unweighted consumption they ignore the well-established concept in economics of diminishing marginal value of consumption; second, they provide no information about the social distribution of costs and benefits and their impact on inequalities (see discussion in Cookson et al. (2020)). There is a large literature on the theoretical and practical shortcomings of unweighted cost-benefit analysis and the advantages of alternative utilitarian and prioritarian approaches to economic evaluation based on explicit individual wellbeing and social welfare functions (Adler and Fleurbaey, 2016).

Our framework generates individual-level outcomes that could be used in many different ways to create summary indices of wellbeing for use in economic evaluation. In our illustrative evaluation we follow Cookson et al. (2020) who propose a simple approach based on the quality-adjusted life year (QALY) concept in health economics but adjusting for consumption as well as health-related quality of life. Our approach could be used to construct many other multidimensional measures of wellbeing that have been proposed in the literature, including equivalent income measures and measured based on life satisfaction (Adler and Fleurbaey, 2016). Cookson et al. (2020) refer to their approach as an “equivalent life” approach (Canning, 2013), and the resulting wellbeing metric as “years of good life” or “wellbeing QALYs”. Following them, we represent individual wellbeing in year *t* by a function *w*_*t*_() increasing in both consumption and health. More specifically, *w*(..) = *health*_*i,age*_ + *u*(*consumption*_*i,age*_) where *u*(.) is a standard isoelastic utility of income function defined as 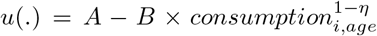. The parameter *η >* 1 captures diminishing marginal value of income, and *A* and *B* are constants which depend on normative parameters: *η* (already mentioned), minimal consumption for a life worth living and standard consumption for a good life. In the current application we set minimal consumption at £1,000 (estimated amount required to buy basic food supplies in the UK for a year) and standard consumption at £24,000 (the mean consumption in the LifeSim simulated cohort), and *η* = 1.26 (see Cookson et al. (2020)).

The interpretation is that a good year is a year lived enjoying full health and consuming the equivalent of the average consumption in a rich country. The good-years measure is more informative than conventional monetary measures because it takes into account the notion that one pound of additional consumption is worth substantially more to a poor individual than a rich individual.

### 2.5 Computing Methods

LifeSim is implemented in software R (tested on R version 3.6.2) using object-oriented programming for R (requires R6 and tidyverse packages). The code and related data files compressed in a zip-file ‘LifeSim.zip’ can be extracted and run on a high performance computing (HPC) cluster (Slurm Workload Manager).

When we split the simulation into 500 partitions, it takes 28 minutes to run it on the HPC cluster. The simulation can also be run on a standard PC, for any chosen number of individuals.

The code is written using an object-oriented approach built around individuals, capturing their initial endowments and the skills and assets they acquire through life as they undergo various experiences, the probability of which are influenced by their past histories. This allows us to simulate individual life histories in an intuitive manner and easily communicate and validate our modelling assumptions in discussion with domain experts in various stages of the life-course. The code is currently written in R allowing us to elegantly incorporate advanced statistical methods into our modelling. However, R being an interpreted language can be slow to run and if performance was a concern our code could easily be translated into a compiled object oriented programming language such as C++. There are also ways of re-writing the original R code in more compact ways, known as “vectorisation”, which are harder for non-specialists to follow but faster to run because they avoid conventional programming loops that require the same time-consuming interpretation operations to be applied repeatedly.

## 3 Baseline Results

In this section we show our baseline simulation results, and demonstrate some formats in which they can be analysed.

Table 3 provides key summary statistics for the simulated outcomes, including child outcomes, adult outcomes and final wellbeing outcomes. We show means, standard deviations, and the minimum and maximum value of an outcome in the total distribution of the simulated individuals in the baseline simulation, as well as means and standard errors for a bootstrap simulation, i.e. after running the simulation 100 times with a different random seed each time. Table 3 does not present the summary statistics of the the initial conditions, as well as the child’s cognitive skills and SDQ scores that we obtain from the childhood survey dataset (MCS), as these variables have already been summarised in Table 1.

**Table 3:** Summary Statistics of the Simulated Outcomes.

| Outcome | Baseline simulation |  |  |  | Bootstrap simulation |  |
| --- | --- | --- | --- | --- | --- | --- |
|  | Mean | SD | Min | Max | Mean | SE |
| <u>Child outcomes</u> |  |  |  |  |  |  |
| Conduct disorder at age 5, % | 8.63 | 28.08 |  |  | 8.59 | 0.10 |
| Conduct disorder at age 18, % | 9.09 | 28.74 |  |  | 9.00 | 0.08 |
| <u>Adult outcomes</u> |  |  |  |  |  |  |
| Proportion of university graduates, % | 38.51 | 48.66 |  |  | 38.51 | 0.15 |
| Proportion of working years in unemployment, % | 5.73 | 6.63 | 0.00 | 100.00 | 5.70 | 0.02 |
| Proportion of lifetime in poverty, % | 26.85 | 18.30 | 0.00 | 100.00 | 26.80 | 0.05 |
| Proportion of working years in prison, % | 1.62 | 5.12 | 0.00 | 75.00 | 1.60 | 0.01 |
| Proportion of retirement in residential care, % | 1.28 | 3.95 | 0.00 | 100.00 | 1.28 | 0.02 |
| Proportion of adult years as a smoker, % | 5.32 | 5.12 | 0.00 | 100.00 | 5.31 | 0.02 |
| Proportion of adult years with CHD, % | 6.16 | 4.07 | 0.00 | 33.33 | 6.15 | 0.01 |
| Proportion of life years with mental illness, % | 5.98 | 2.81 | 0.00 | 25.81 | 5.99 | 0.01 |
| Years of life | 78.78 | 13.04 | 0.00 | 100.00 | 78.80 | 0.04 |
| Premature mortality rate (before age 75), % | 28.13 | 44.96 |  |  | 28.04 | 0.14 |
| Annual earnings (lifetime average), GBP | 29,655 | 7,638 | 4,792 | 67,879 | 29,659 | 23 |
| Annual savings (lifetime average), GBP | 2,833 | 942 | 0 | 7,803 | 2,832 | 3 |
| Annual interest (lifetime average), GBP | 402 | 234 | 0 | 3,321 | 402 | 1 |
| <u>Final wellbeing outcomes</u> |  |  |  |  |  |  |
| Annual consumption (lifetime average), GBP | 24,114 | 6,648 | 10,000 | 113,817 | 24,115 | 22 |
| Healthy years | 68.28 | 9.99 | 0.87 | 88.16 | 68.30 | 0.03 |
| Healthy years (discounted) | 40.94 | 3.96 | 0.87 | 48.01 | 40.96 | 0.01 |
| Good years | 65.67 | 10.21 | 0.69 | 91.93 | 65.69 | 0.03 |
| Good years (discounted) | 39.80 | 4.89 | 0.69 | 52.18 | 39.82 | 0.01 |

Note: The baseline simulation mean, standard deviation (SD), minimum value (Min) and maximum value (Max) are calculated for the simulated population of 100,000 (for the lifetime aggregates, or yearly – for the annual variables). The bootstrap simulation mean and standard error (SE) are calculated for the distribution of the means of the 100 bootstrap simulations. The time periods for calculating life-stage proportions are as follows: ‘working years’ refer to the period between ages 19-69; ‘retirement’ refers to the time period from age 70 up to death; adult years refer to the time period from age 19 up to death; lifetime refers to the entire period from birth to death. CHD – coronary heart disease. We use year 2015/16 prices and the annual discount rate of 1.5% (Paulden and Claxton, 2012).

The baseline simulation means do not differ much from the bootstrap means, and the bootstap standard errors are small, implying that changing the random seed has a negligible effect on the simulated outcome means with the simulation size that we use.

Approximately 9% of 18 year-old adults develop conduct disorder in the LifeSim simulation. This estimate fits within the range of 1-10 %, commonly reported in the epidemiology literature on conduct disorder (see a review in Hinshaw and Lee (2003), also Patel et al. (2018)). Our estimate, however, slightly exceeds the 8% of young men and 5% of young women with conduct disorder estimated by Mental Health of Children and Young People in England survey in year 2017. This small difference may be caused by the fact that the algorithm by Goodman et al. (2003); Goodman, Renfrew and Mullick (2000) that we use to simulate conduct disorder incidence is based and validated on child samples attending child mental health clinics, and it may overestimate the actual conduct disorder prevalence in the general population. On the other hand, conduct disorder diagnosis in the clinic sample can be argued to be more precise and sensitive than in the survey data sample, because in the clinic sample diagnosis was made by mental health specialists using detailed information on symptoms and resultant impairments gathered from multiple informants, whereas in the specific survey sample diagnosis was based on a single specific tool – Development and Well-Being Assessment.

Figure 5 shows the simulated distributions of some core outcomes, which also include the distribution of lifetime wellbeing (measured using the approach by Cookson et al. (2020) described in section 2.4.)

**Figure 5:**
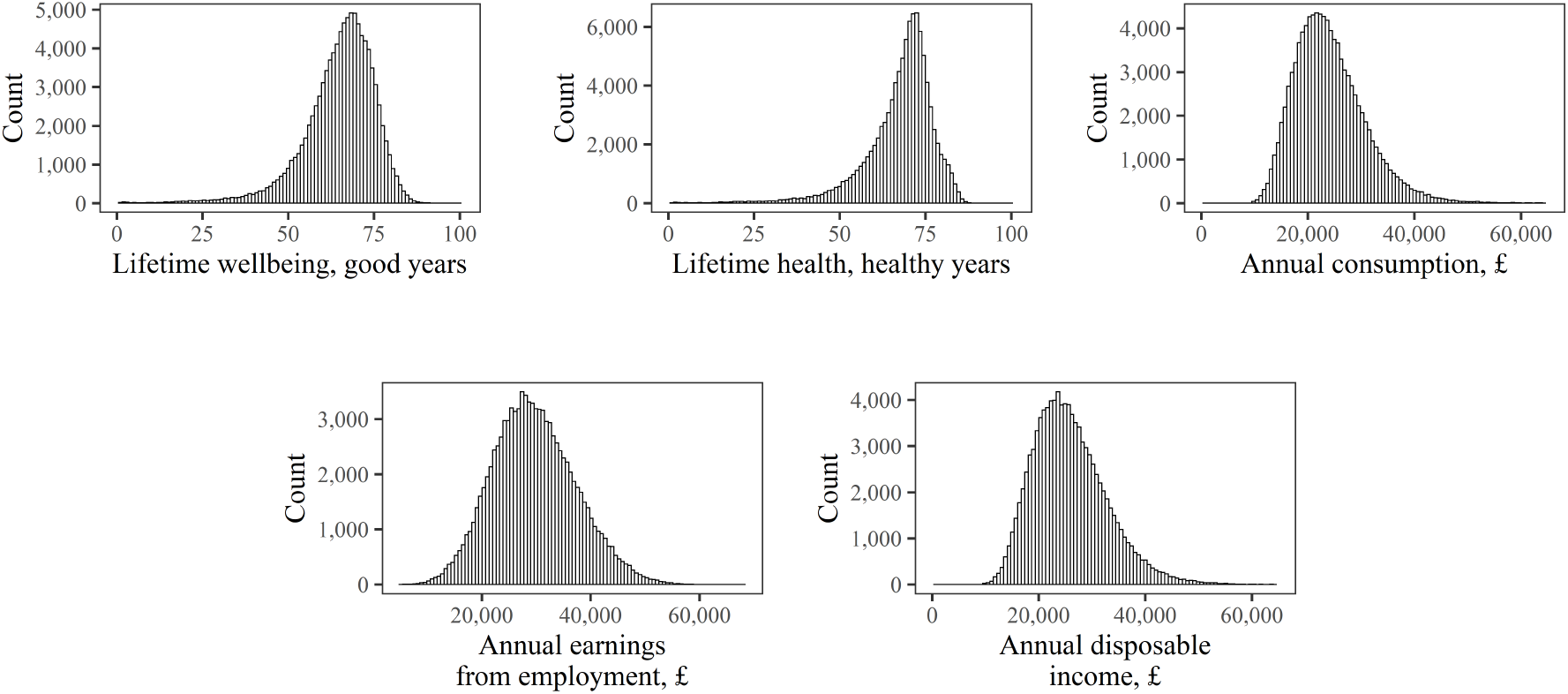
Distributions of Core Outcomes. Note: Disposable income calculated net of taxes, and include earnings from employment, earnings from interest, earnings from pension during retirement years, and cash benefits.

Table 4 shows the average costs to the public budget associated with certain outcomes, cash benefits paid to people who are in poverty or unemployed, as well as taxes on earnings and financial gains. These are calculated over various time intervals over the life-course, and separately for the general population, and then for people born in the lowest and top income quintile groups at birth.^6^

**Table 4:** Cumulative Costs Over Various Time Periods.

| Costs (Per Capita), £ | Age 0-10 | Age 0-15 | Age 0-20 | Age 0-25 | Lifetime |
| --- | --- | --- | --- | --- | --- |
| <u>General Population Cohort</u> |  |  |  |  |  |
| <u>Public Services</u> |  |  |  |  |  |
| Conduct disorder | 1,100 | 1,700 | 1,800 | 1,800 | 1,800 |
| Prison | 0 | 0 | 930 | 3,200 | 17,000 |
| Residential care | 0 | 0 | 0 | 0 | 1,200 |
| <u>Healthcare</u> |  |  |  |  |  |
| CHD | 0 | 0 | 0 | 0 | 1,400 |
| Depression | 740 | 1,900 | 3,600 | 4,900 | 14,000 |
| Other | 9,300 | 14,000 | 20,000 | 27,000 | 88,000 |
| Benefit payments | 1,400 | 2,100 | 4,100 | 6,300 | 12,000 |
| <u>Lowest Income Quintile Group at Birth</u> |  |  |  |  |  |
| <u>Public Services</u> |  |  |  |  |  |
| Conduct disorder | 1,400 | 2,100 | 2,200 | 2,200 | 2,200 |
| Prison | 0 | 0 | 1,100 | 4,000 | 20,000 |
| Residential care | 0 | 0 | 0 | 0 | 1,200 |
| <u>Healthcare</u> |  |  |  |  |  |
| CHD | 0 | 0 | 0 | 0 | 1,400 |
| Depression | 770 | 2,000 | 3,600 | 5,000 | 14,000 |
| Other | 11,000 | 16,000 | 22,000 | 29,000 | 91,000 |
| Benefit payments | 8,600 | 12,000 | 17,000 | 19,000 | 22,000 |
| <u>Top Income Quintile Group at Birth</u> |  |  |  |  |  |
| <u>Public Services</u> |  |  |  |  |  |
| Conduct disorder | 880 | 1,300 | 1,500 | 1,500 | 1,500 |
| Prison | 0 | 0 | 800 | 2,700 | 14,000 |
| Residential care | 0 | 0 | 0 | 0 | 1,100 |
| <u>Healthcare</u> |  |  |  |  |  |
| CHD | 0 | 0 | 0 | 0 | 1,400 |
| Depression | 710 | 1,900 | 3,500 | 4,900 | 14,000 |
| Other | 8,400 | 13,000 | 18,000 | 25,000 | 85,000 |
| Benefit payments | 0 | 0 | 1,300 | 3,000 | 7,900 |

Note: All values are calculated per simulated individual in year 2015/16 prices, and discounted at 1.5% annual rate, and rounded to 2 significant figures. See details on cost sources in table A.6 in Appendix A.

Table 5 provides two summary measures of inequality, based on differences in lifetime expected wellbeing between best off and worst off groups on the basis of the following early childhood circumstances – sex, parental income quintile group (poorest vs. richest 20%), parental mental health, parental education, and high baseline conduct problems (SDQ conduct problem score at age 5 equal to 7 or above). Our “extreme best off group” focuses on individuals in the top category of all four main markers of social disadvantage in early life (top 20% parental income, high parental education, no parental mental illness, high baseline conduct problems). Our “best off 20% group” focuses on the best off 20% of individuals in terms of predicted lifetime wellbeing based on all four main markers of social disadvantage in early life.

**Table 5:**
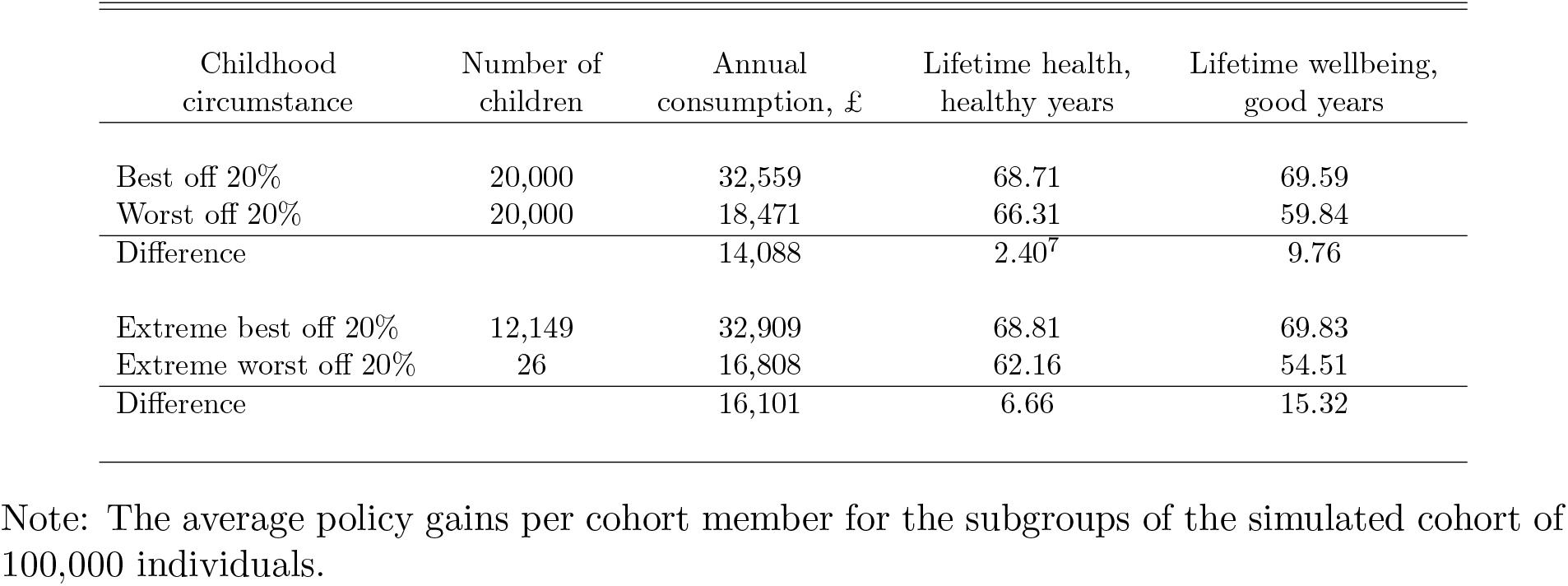
Whole Cohort Lifetime Inequality by Childhood Circumstance.

| Childhood circumstance | Number of children | Annual consumption, £ | Lifetime health, healthy years | Lifetime wellbeing, good years |
| --- | --- | --- | --- | --- |
| Best off 20% | 20,000 | 32,559 | 68.71 | 69.59 |
| Worst off 20% | 20,000 | 18,471 | 66.31 | 59.84 |
| Difference |  | 14,088 | 2.40<sup>7</sup> | 9.76 |
| Extreme best off 20% | 12,149 | 32,909 | 68.81 | 69.83 |
| Extreme worst off 20% | 26 | 16,808 | 62.16 | 54.51 |
| Difference |  | 16,101 | 6.66 | 15.32 |

Note: The average policy gains per cohort member for the subgroups of the simulated cohort of 100,000 individuals.

## 4 Comparison With Other Datasets

### 4.1 Comparison With 1970 Birth Cohort Study

Table 6 compares the LifeSim predictions with data from the 1970 Birth Cohort Study (BCS70) at ages 26, 29, 42 and 46, as a simple validation check. We list the number of observations, means and standard-deviations of the LifeSim variables for children born in the year 2000 and the BCS70 variables for children born in the year 1970, representing the same outcomes. For each outcome, we quantify the difference between the LifeSim distribution and BCS70 distribution in terms of the absolute difference in their means and standard deviations.

**Table 6:** Comparison with the British Cohort 1970.

| Outcome | N |  | Mean |  | Difference<br>in means | SD |  | Difference<br>in SDs |
| --- | --- | --- | --- | --- | --- | --- | --- | --- |
|  | LifeSim | BCS70 | LifeSim | BCS70 |  | LifeSim | BCS70 |  |
| Age 26 |  |  |  |  |  |  |  |  |
| Male (indicator) | 99,402 | 9,003 | 0.48 | 0.46 | 0.03 | 0.50 | 0.50 | 0.00 |
| University Degree (indicator) | 99,402 | 8,399 | 0.39 | 0.25 | 0.13 | 0.49 | 0.43 | 0.05 |
| Employed (indicator) | 99,402 | 9,003 | 0.95 | 0.96 | -0.01 | 0.23 | 0.20 | 0.02 |
| Earnings (in year 2015 £) | 99,402 | 6,642 | 19,736 | 14,279 | 5,457 | 5,882 | 7,110 | -1,228 |
| Depression (indicator) | 99,402 | 9,003 | 0.07 | 0.10 | -0.03 | 0.25 | 0.30 | -0.05 |
| Smoking (indicator) | 99,402 | 8,892 | 0.06 | 0.27 | -0.20 | 0.25 | 0.44 | -0.20 |
| Age 29 |  |  |  |  |  |  |  |  |
| Male (indicator) | 99,267 | 11,261 | 0.48 | 0.49 | -0.00 | 0.50 | 0.50 | -0.00 |
| University Degree (indicator) | 99,267 | 11,211 | 0.39 | 0.27 | 0.11 | 0.49 | 0.44 | 0.04 |
| Employed (indicator) | 99,267 | 9,506 | 0.94 | 0.96 | -0.02 | 0.23 | 0.19 | 0.04 |
| Earnings (in year 2015 £) | 99,267 | 8,102 | 22,400 | 20,796 | 1,604 | 6,682 | 68,798 | -62,115 |
| Depression (indicator) | 99,267 | 11,261 | 0.07 | 0.10 | -0.03 | 0.25 | 0.30 | -0.05 |
| Smoking (indicator) | 99,267 | 11,205 | 0.06 | 0.29 | -0.23 | 0.24 | 0.45 | -0.22 |
| Age 42 |  |  |  |  |  |  |  |  |
| Male (indicator) | 98,149 | 9,841 | 0.48 | 0.48 | 0.00 | 0.50 | 0.50 | 0.00 |
| University Degree (indicator) | 98,149 | 9,841 | 0.39 | 0.34 | 0.05 | 0.49 | 0.47 | 0.01 |
| Employed (indicator) | 98,149 | 8,594 | 0.95 | 0.97 | -0.02 | 0.21 | 0.16 | 0.05 |
| Earnings (in year 2015 £) | 98,149 | 2,158 | 29,327 | 22,107 | 7,220 | 9,604 | 15,567 | -5,963 |
| Depression (indicator) | 98,149 | 9,756 | 0.07 | 0.11 | -0.04 | 0.26 | 0.31 | -0.05 |
| Smoking (indicator) | 98,149 | 9,801 | 0.06 | 0.20 | -0.14 | 0.24 | 0.40 | -0.17 |
| Age 46 |  |  |  |  |  |  |  |  |
| Male (indicator) | 97,532 | 8,581 | 0.48 | 0.48 | -0.00 | 0.50 | 0.50 | -0.00 |
| University Degree (indicator) | 97,532 | 8,444 | 0.39 | 0.34 | 0.04 | 0.49 | 0.47 | 0.01 |
| Employed (indicator) | 97,532 | 5,038 | 0.95 | 0.99 | -0.03 | 0.21 | 0.12 | 0.09 |
| Earnings (in year 2015 £) | 97,532 | 358 | 28,558 | 22,538 | 6,020 | 9,978 | 31,559 | -21,581 |
| CHD (indicator) | 97,532 | 8,353 | 0.02 | 0.00 | 0.02 | 0.15 | 0.05 | 0.10 |
| Depression (indicator) | 97,532 | 8,486 | 0.06 | 0.14 | -0.08 | 0.23 | 0.35 | -0.12 |
| Smoking (indicator) | 97,532 | 8,578 | 0.05 | 0.15 | -0.10 | 0.23 | 0.36 | -0.13 |

Note: N – number of observations, SD – standard-deviation. We quantify the difference between the LifeSim distribution and BCS70 distribution in terms of the absolute difference in their means and standard deviations. Earnings is the net pay from employment.

We would expect some adult outcomes to be similar (e.g. health) but others to be substantially different (e.g. earnings, rates of smoking and university education), and so this can be seen as a simple validation check to ensure that our model provides broadly similar findings in the same ballpark where appropriate, and substantially different findings where we know different generations had very different experiences e.g. smoking. Nevertheless, most variables do not deviate substantially from the same quantities characterising the cohort born in 1970.

One exception already mentioned is smoking, which is expected and can be explained by the change in smoking rates over time. Another exception is education – the proportion of people with a degree under 30 years old – which is much higher in the LifeSim cohort. This can be explained by the change in higher education participation rates over time, and increased equality between the genders in the cohort born in 2000. Over time the 1970s cohort partially catches up with the LifeSim cohort by obtaining qualifications at a later age – at the age 46 the proportion of people with a university degree is more similar in both samples than at the age 26. Finally, the LifeSim earnings at all ages on average exceed the 1970s cohort earnings. This can be explained by cohort effects, such as general differences in economy, society, culture and politics experienced by the two cohorts.

### 4.2 Comparison With Recent Cross-Sectional Data

To avoid such general cohort effects which arise when comparing two generations born 30 years apart, we also carry out a simple validity check using more recent cross-sectional datasets. More specifically, we compare our age-specific LifeSim outcomes with age-specific outcomes in cross-sectional data.

Figure 6 compares the age-earnings profile for males and females in the LifeSim simulation with our target dataset – ONS Annual Survey of Hours and Earnings in year 2015, and in the Understanding Society survey in year 2015. The concave trend with age, initially increasing and then – decreasing earnings, is very similar in the tree datasets.

**Figure 6:**
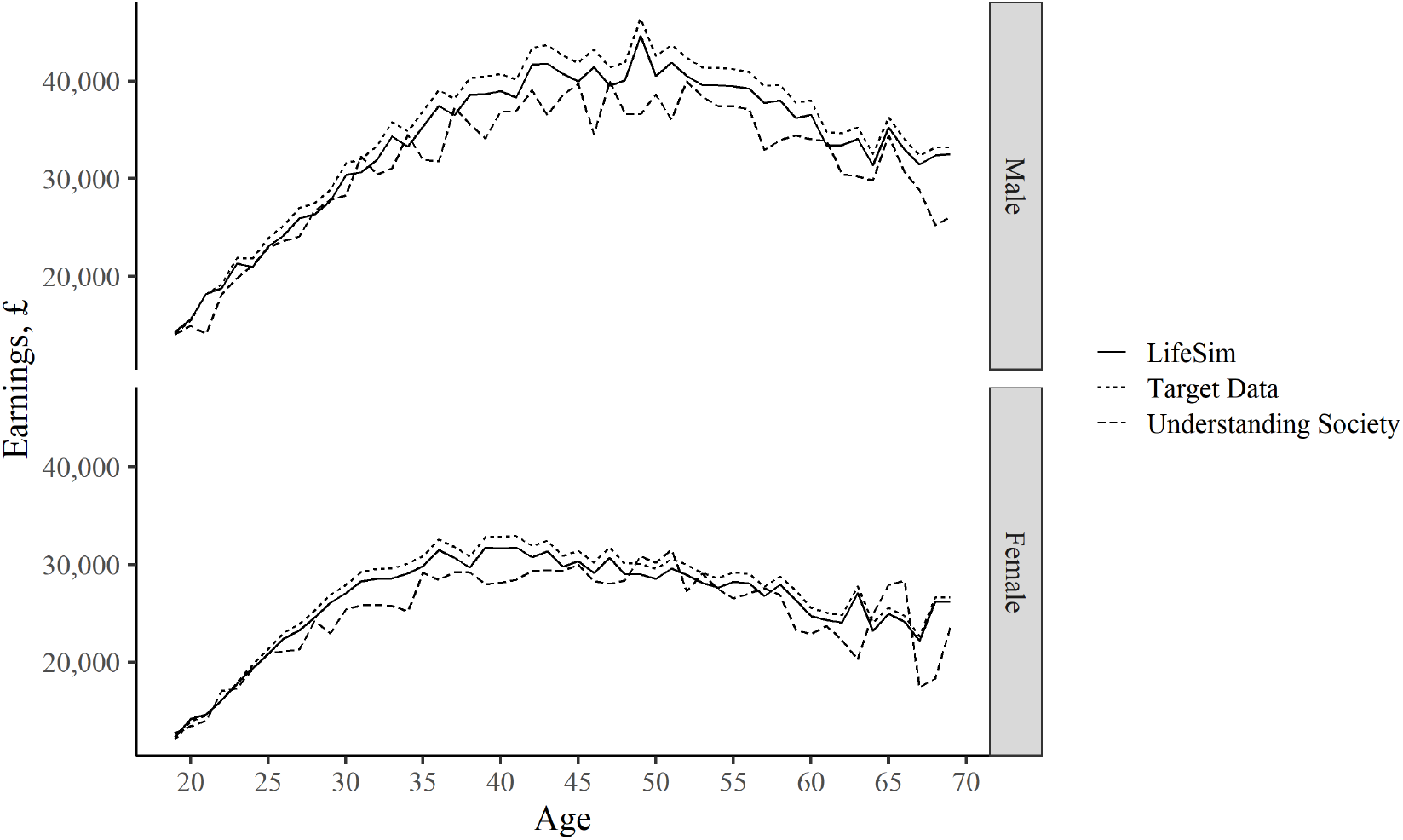
Earnings: Comparison with Other Data Sources. Note: Target data is the annual full-time gross pay data from Annual Survey of Hours and Earnings, ONS, 2015. See more details in table A.2. The Understanding Society data is the estimated annual full-time gross pay data from wave 7 (started in year 2015), calculated for individuals in full-time employment as usual gross pay times the frequency of pay per year. All earnings are in year 2015 £.

Figure 7 compares the earnings distributions by sex and different age groups in the LifeSim cohort and the Understanding Society data. Both distributions have similar medians for the different sex-age groups, and also become more uniform with increasing age. One issue left to be addressed as part of future work is modelling of the relatively longer right hand side tail which can be observed for the Understanding Society data and not for the LifeSim data. This tail represents the highest-earning people in the distribution. The LifeSim earnings output does not have this tail, as we do not model the outcome of being employed in extremely-high earning jobs. Addressing this feature in LifeSim would require modelling the link with variables in early life that would lead to such extremely-high earning states.

**Figure 7:**
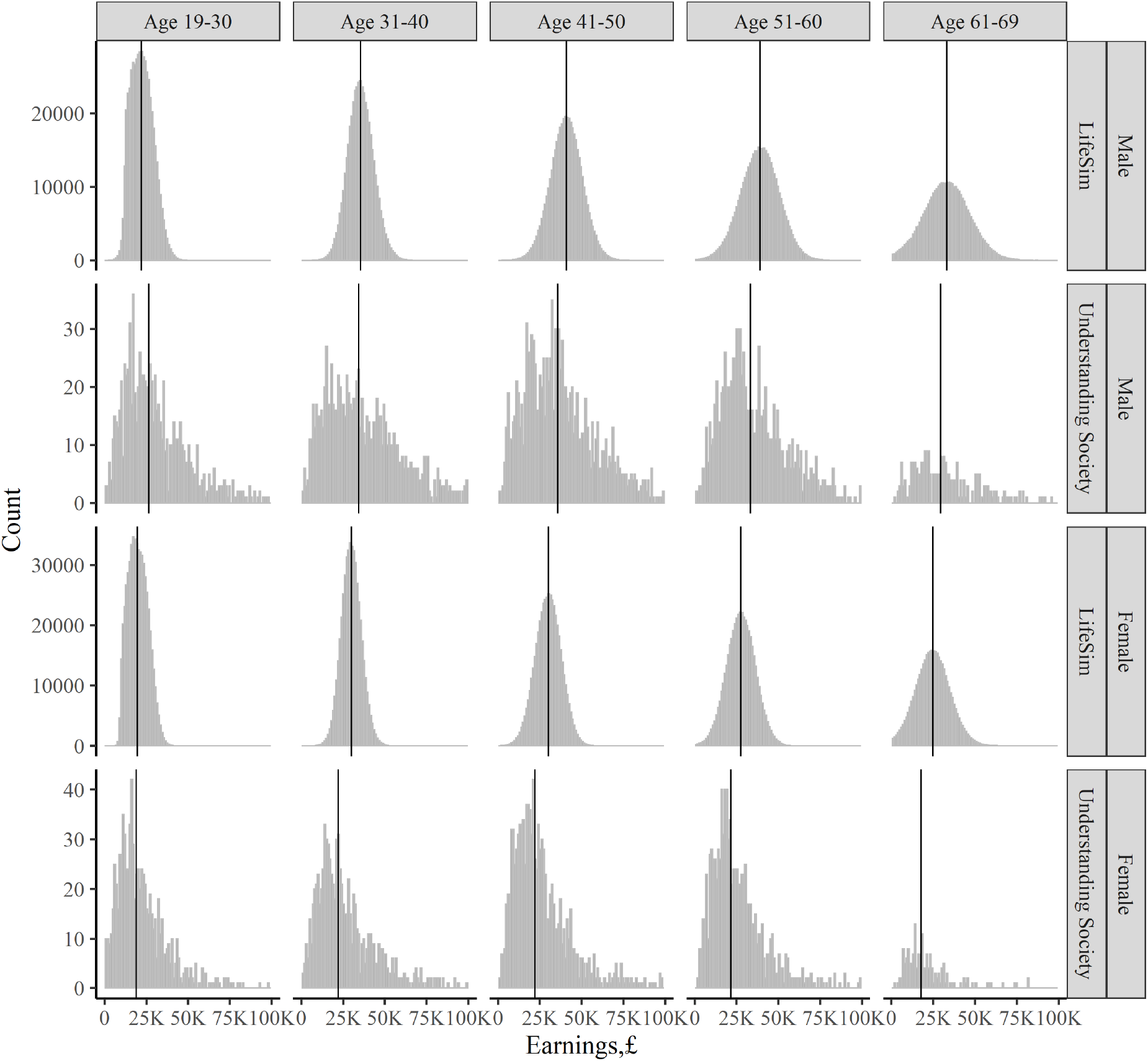
Other Outcomes: Comparison with Understanding Society. Note: Data is the estimated annual full-time gross pay data from Understanding Society]wave 7 (started in year 2015), calculated for individuals in full-time employment as usual gross pay times the frequency of pay per year. All earnings in year 2015 £. The black lines denote medians.

In Figure 8 we compare the prevalence of the different discrete outcomes in LifeSim cohort, and in our corresponding target datasets, which include Health Survey for England for the health-related outcomes, ONS Labour Force Survey for unemployment and Department for Education estimates for participation in higher education.

**Figure 8:**
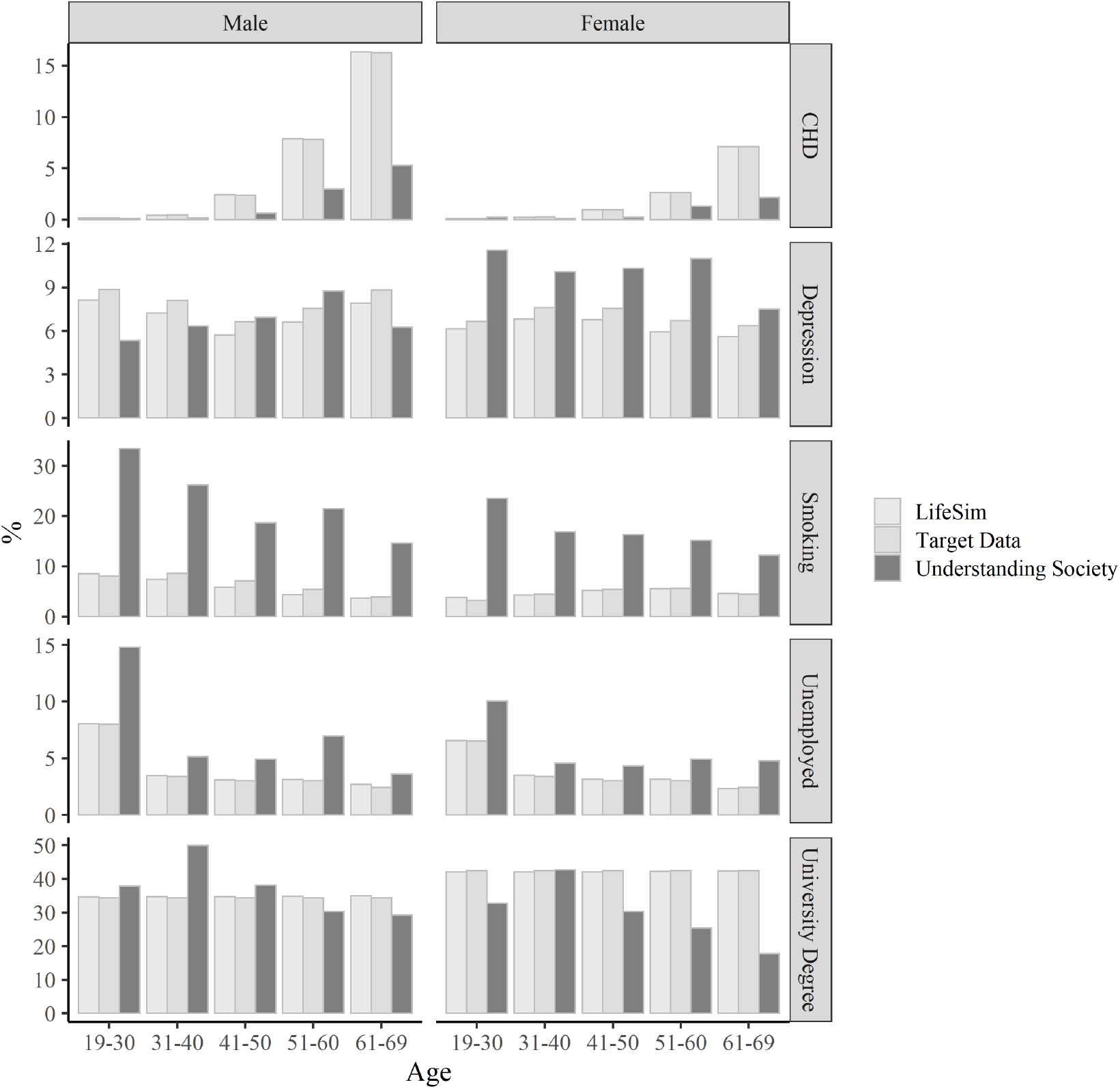
Earnings: Comparison with Other Data Sources. Note: Target data for the outcomes is as follows. Depression – Health Survey for England, 2014; CHD – Health Survey for England, 2006; smoking– Health Survey for England, 2006; employment – Labour Force Survey, ONS, 2018; university degree – Department for Education, 2016.

The simulated outcomes matches the target data well, but there is some small discrepancy with the Understanding Society data, which can be explained by differences how data on similar outcomes is collected across different surveys.

## 5 Discussion

Microsimulation offers a forward-looking alternative to conventional approaches to long-term childhood policy analysis, which have often relied on long-term follow-up of high-profile historical childhood policy experiments that took place decades ago – such as the US Abecedarian experiment (García et al., 2020) – and are of questionable relevance to current policy decisions. We present LifeSim – a proof of concept microsimulation model for analysing the full long-term consequences of childhood policies from a lifetime perspective. LifeSim is capable of modelling a rich set of developmental, social, economic and health outcomes from birth to death for each child in a general population birth cohort of 100,000 English children born in the year 2000-1, together with public costs and summary wellbeing metrics.

Since our model is designed for the purpose of partial equilibrium policy analysis rather than forecasting of macro-level trends, the most important criteria for model credibility arguably relate to the quality of the underlying conceptual framework and data sources rather than ability to predict population-level trends (Kopec et al., 2010). Nevertheless, we provide a simple comparison of our simulation with external data on population-level trends. First, we provide a comparison with data from the 1970 Birth Cohort Study up to age 46. We find that our simulation is broadly consistent with the external data and substantially divergent when appropriate – for example, our simulation for people born in 2000 has a much lower proportion of people smoking than the 1970 cohort, reflecting the reduction in smoking rates in the UK since the 1970s. Also, our simulation for people born in 2000 has a much larger proportion with young people having obtained a university degree at age 26 than the 1970 cohort at that age, reflecting the massive expansion in university provision in the UK since the 1970s.

We also provide a comparison with a recent external cross-sectional dataset – Understanding Society (in the year 2016). Our simulated earnings outcome replicates reasonably well the sex-age specific distributions observed in the Understanding Society data. Also, for our simulated key discrete outcomes – including health-related outcomes and unemployment – the sex specific prevalence trends against age are not too deviant from the trends observed in the Understanding Society data. Any minor discrepancies can be explained by differences in data collection methods for Understanding Society and our target datasets.

Finally, we provide an additional check of LifeSim output against the various target datasets that we directly use to calibrate our equations, such as Health Survey for England, and Office for National Statistics datasets. As expected, our simulated outcomes match very well the trends and patterns observed in the target data. Because our model is flexible and can be used together with many data sources, if needed, one can easily substitute our target datasets with alternative datasets, to match the trends and patterns observed in these alternative sources.

The main strength of our model is that it captures the dynamic individual-level interaction between many outcomes across the social, economic and health domains over the entire lifecourse. Previous models have modelled either two or three of these domains or only a part of the lifecourse. Simultaneously analysing many outcomes allows us to capture how many early life disadvantages can compound over the lifecourse creating a spiral of multiple disadvantage.

Another strength of LifeSim is that it simulates the long-run outcomes for a whole general population cohort of children, not just analysing the outcomes of a narrow group of trial participants. This allows carrying out more complex and policy-relevant analysis, including assessment of the distributional impacts and policy opportunity costs on the general population, and exploring options for targeting the policy to different subgroups of the population.

LifeSim also generates long-term individual-level data, which makes it compatible with applying new multidimensional summary indices of wellbeing recently proposed in the theoretical literature (Cookson et al., 2020; O’Donnell et al., 2014; Fleurbaey et al., 2013; Fleurbaey and Schokkaert, 2013). These indices are more informative than conventional monetary valuation based on aggregate outcomes, as they allow to account for the diminishing marginal value of consumption and other sources of heterogeneity in the marginal value of different life outcomes to different individuals. However, application of these indices in practice requires individual level long-term time series data on many outcomes across the health, social and economic outcome domains. Such rich long-term data is difficult to obtain from existing datasets, especially if we are interested in analysing cohorts living in present rather than historical cohorts of people born decades ago. Models such as LifeSim can compile the many data sources together to extrapolate the required individual-level long term outcomes.

Perhaps the most important limitation of our modelling approach is the assumption that micro-level causal pathways are invariant to social trends and policy intervention. LifeSim can readily accommodate macro-level social trends, such as changes in average earnings and educational attainment, by using up-to-date target data. However, some social trends do raise potential threats to our fundamental assumption of causal pathway invariance. For example, the massive expansion in higher education participation since the early 1990s means that the “signalling” value of a university degree has diminished as a way of helping employers to identify job candidates with exceptional ability. The proportional effect of obtaining a university degree on adult earnings for the 2000 cohort graduating in the early 2020s is thus likely to be smaller than the effect estimated in the 1970 cohort graduating in the early 1990s. We do not take into account potentially measurable biases of this kind in the effect estimates used in the present version of LifeSim, which are based on existing published analysis of longitudinal data on historical cohorts of children. However, this limitation could be addressed in future work by identifying the most important potential biases in effect estimates from longitudinal data on historical cohorts and making appropriate adjustments through careful synthesis of evidence and elicitation of expert opinion. Relatedly, specific transition pathway estimates could also be modified in specific cases to strengthen external validity for specific populations. For example, estimates based on long-term outcomes for mostly white children born in the 1970s may not be applicable to Asian British populations. Using external data sources to estimate long-run health effects for Asian British populations would produce more applicable estimates for those populations.

Further, in principle, using up-to-date cross-sectional target data as well as effect estimates is a methodological strength of our approach to parameterisation, which can help to improve the external validity of the model by correcting for omitted variable and cohort biases in our effect estimates and ensure that our predictions are calibrated in line with current population-level outomes. However, the current version of our model relies more heavily on cross-sectional target data than effect estimates, which is a limitatation from the perspective of internal validity and causal inference. This is an important limitation, given the intended use of the model for the purpose of policy analysis and evaluation. Future work in developing a version of the model for routine policy analysis could aim to reverse this balance, placing more emphasis on credible effect estimates to improve internal validity while maintaining a role for target data to ensure external validity.

LifeSim can also be extended to incorporate additional features. One extension would be to incorporate more outcomes. Our model includes many different categories of human capital (e.g. cognitive skills, social skills, educational attainment, health, employment) but within each category, more nuanced distinctions could be made. Health outcomes are modelled using just three binary variables – mental illness (depression), physical illness (CHD) and mortality – educational outcomes focus only on gaining a university degree; employment outcomes focus only on unemployment not precarious employment; and our modelling of the tax and benefit system and retirement savings is extremely stylised. Similarly, more individual-level factors could be included (e.g. ethnicity), more family-level factors (e.g. child abuse) and it would also be possible to add neighbourhood-level factors (e.g. neighbourhood-level deprivation in childhood, as well as family-level income). Also, our tax benefit modelling is highly stylised and could be improved by incorporating a standard static tax benefit calculator, such as Euromod (Sutherland and Figari, 2013).

LifeSim also currently does not model many important outcomes during childhood, such as cognitive skills, but rather takes them as given from the MCS. Future work could undertake formal dynamic modelling of all the relevant outcomes during childhood and adolescence, based on structural equation modelling and mediation analysis of MCS data that estimates all the relevant parameters in a single, integrated longitudinal data analysis.

Next, LifeSim parameters that govern the evolution of lifecourse outcomes during different stages of childhood and adulthood are currently estimated separately using different studies based on different datasets, specifications and estimation methods. This increases the parameter uncertainty, which compounds over time when modelling lifecourse trajectories. Future childhood policy modelling could adopt a more joined-up and systematic approach to estimate these parameters simultaneously by linking together data on different stages of the lifecourse from successive cohort studies (Hughes, Tilling and Lawlor, 2020). This would make the model more ready for prime time policy analysis, and also allow a formal analysis of parameter uncertainty by bootstrapping parameters using an estimated variance-covariance matrix.

Another extension would be to re-calibrate our model to other populations – e.g. the UK in 2025, or England or Scotland, or a sub-national area of England – by updating the initial conditions of the birth population and the external macro target data on average population level outcomes and associations within that birth population in subsequent years.

Furthermore, LifeSim currently does not model interactions between individuals, and an important extension would be to model interactions, such as the dynamics of family formation and dissolution and spillover effects on other family members. Building an interactive model would also allow modeling the effects of infectious disease transmission, as well as the non-communicable mental and physical illnesses that are currently the focus of the model.

Our model structure could also be extended in more fundamental ways – for example, to model the all-age population rather than just a birth cohort, and to model parental investment choices and other behavioural responses that may depend on social trends, changes in the policy environment, and/or the behaviour of other individuals. It should be acknowledged that considering any extensions involves making trade-offs between model complexity and tractability, and in some cases it may be preferable to use other more specialist models and combine the findings from different models, rather than expand an existing model. For example, as already mentioned – our model could be combined with Euromod (Sutherland and Figari, 2013) – the tax and benefit microsimulation model, to generate more comprehensive output on taxes and benefits for the assessment of the consequences to the public budget.

Overall, LifeSim is a flexible childhood policy model which serves as proof of concept in demonstrating the potential added value of lifecourse microsimulation in long-term childhood policy analysis. It sets a foundation for the development of a long-term childhood policy model which can be routinely used to carry out prime time policy analysis.

## Data Availability

All the data referred to in the manuscript is publicly available data.

## Acknowledgements

We would first like to thank the members of our advisory group: Annalisa Belloni, Sarah Cattan, Leon Feinstein, Paul Frijters, Peter Goldblatt, Heather Joshi, Catherine Law, Lara McClure and Christine Power.

For useful comments we also are grateful to Shehzad Ali, Mark Ashworth, Karen Bloor, Laura Bojke, Eva Maria Bonin, Jonathan Bradshaw, Penny Breeze, Alan Brennan, Eric Brunner, Tracey Bywater, Simon Capewell, Maria Guzman Castillo, Bette Chambers, Brendan Collins, Gabriella Conti, Peter Diggle, Tim Doran, Susan Griffin, Nils Gutacker, James Heckman, Nathan Hendron, Bruce Hollingsworth, Andrew Jones, Noemi Kreif, Christodoulos Kypridemos, Richard Mattock, Cheti Nicoletti, Owen O’Donnell, Martin O’Flaherty, Kate Pickett, George Ploubidis, Gerry Richardson, Jemimah Ride, Matthew Robson, Tracey Sach, Filipa Sampaio, Trevor Sheldon, Tushar Srivastava, Mark Strong, David Taylor-Robinson, Valentina Tonei, Aki Tsuchiya, Simon Walker, Margaret Whitehead and Mark Mon Williams, and anonymous reviewers of previous versions of the manuscript.

We would also like to thank Matteo Ricciardi and two anonymous reviewers for detailed and constructive comments on our original submission to the International Journal of Microsimulation.

The errors and opinions expressed in this paper are our own.

## Footnotes

1 Internal validity relates to claims about cause and effect within the study population, whereas external validity relates to how applicable the findings are to real world policy settings.

2 According to Statistics Canada, they developed a dynamic microsimulation model in the 1990s with a rich set of co-evolving economic, social and health outcomes, called LifePaths (Spelauer et al., 2013), which has subsequently been discontinued. However, this model seems to have had limited detail on developmental outcomes in childhood and we could not find detailed technical information or any published economic evaluations based on it.

3 Because the MCS data was only available up to age 14 when our model was developed, we approximate the outcomes between ages 15-18 using the MSC data for age 14; this can now be updated with the data from the latest MCS wave which has recently become available and at which children are 17 years old.

4 More specifically, the algorithm allocates a probability of 0.61 for children with the SDQ conduct problem score of at least 5 combined with the impact score of at least 2; a probability of 0.31 for children with the conduct problem score equal to 4 (irrespective of the impact score) and a probability of 0.06 for all the other children with conduct problem scores below 4.

5 This equation and other equations in this section are simplified examples of the actual equations that we use; see Appendix A for the full mortality equation and the other equations that we use.

6 The total residential care cost figure does not include the substantial private costs of residential care, which we assume fall on individuals if they have sufficient savings, nor the public costs of residential care before the age of 60. It may be an underestimate of public costs, because we make simple and conservative assumptions about the need for residential care and eligibility for public funding - for example, we use simple sex-specific rates of care home use in people aged 65 and over (2% for men and 4% for women) but do not model the rapid age-related increase in risk which results in much higher rates for people surviving into their 80s and belyond.

7 Standard period estimates of gaps in healthy life expectancy by current socioeconomic status are substantially larger than our cohort estimate of gaps by early childhood circumstance, due to dynamic interdependence between health and social status over the lifecourse. Adult-onset illness that is unrelated to early childhood circumstances may cause downward social mobility, and deterioration of social and economic outcomes that is unrelated to early childhood circumstances may cause deteriorating health.

