## Appendix for "LifeSim: A Lifecourse Dynamic Microsimulation Model of the Millennium Birth Cohort in England"

### Supplementary Appendices for "LifeSim: A Lifecourse Dynamic Microsimulation Model of the Millenium Birth Cohort in England"

Ieva Skarda, Miqdad Asaria and Richard Cookson

**Appendix B References for Data and Parameter Sources** **20**

#### Appendix A Modelling Equations

This appendix summarises the principles that we follow and data that we use to build our modelling equations.

##### A.1 Functional Forms

The functional form of each modelling equation is chosen depending on (a) the type of variable that we model (continuous quantity vs. indicator) and (b) the format in which the parameter estimate is reported (e.g. coefficient estimates from a linear regression, odds ratios from a logistic regression, percentage changes, etc.).

###### A.1.1 Modelling A Continuous Quantity

Literature on continuous quantities (e.g. earnings, IQ scores, age at death) most often report parameter estimates in the form of beta coefficients from a linear regression, that represent either (i) absolute change in the independent variable (Y) as a result of a unit change in the dependent variable (X); or (ii) relative (or percentage) change in Y, as a result of a unit change in X. Following this, we model the effects on Y either as absolute or relative changes.

**Absolute Change.** Assume that we want to model $\beta_{cog}^{earnings}$ – the linear effect of individual cognitive ability at age 18 (denoted $cog_{i,18}$) on individual earnings at age 19 (denoted $earnings_{i,19}$). The linear specification is:

$$earnings_{i,19} = \alpha_{i,19} + \beta_{cog}^{earnings} cog_{i,18} + error_{i,19} \tag{1}$$

where $\alpha_{i,19}$ captures the constant, as well as the effects of other observable and unobservable variables not explicitly specified in equation (1)); $error_{i,19}$ is random noise with a zero mean.

Equation (1) does not explicitly account for all the possible variables which may drive the term $\alpha_{i,19}$, for example, it does not explicitly model economic conditions, social network and many other characteristics of the modelled individual $i$. To overcome this problem, we assume that the modelled individual $i$ is 'average' in terms of all of the outcomes that we do not explicitly account for, i.e. for all $i = 1,..N$ it holds that $\alpha_{i,19} = \frac{1}{N}\sum_{i=1}^{N}\alpha_{i,19} \equiv \overline{\alpha}[19]$, where $N$ is the number of 19 year-old individuals in the representative population. The term $\overline{\alpha}[19]$ can also be expressed from an averaged equation (1) as $\overline{\alpha}[19] = \overline{earnings}[19] - \beta_{cog}^{earnings}\,\overline{cog}[18]$ and then substituted for $\alpha_{i,19}$ in equation (1), to get:

$$earnings_{i,19} = \overline{earnings}[19] + \beta_{cog}^{earnings} \left(cog_{i,18} - \overline{cog}[18]\right) + error_{i,19} \tag{2}$$

We approximate the average values, such as $\overline{earnings}[age]$, $\overline{cog}[age]$ by averages of survey data representative of the cohort that we model (i.e. what we call 'target data', see Section 3.3 in the main paper and Table A.2 in this Appendix).

**Relative Change.** Assume that a standard deviation increase in the cognitive skills at age 14 is estimated to cause an $k\%$ increase in the earnings at age 19. It can be shown that in this case using the procedure described above will yield:

$$earnings_{i,19} = \overline{earnings}[19] \ \exp\left((1 + \frac{k}{100}) \left(cog_{i,19} - \overline{cog}[14]\right) + error_{i,19}\right) \tag{3}$$

##### A.1.2 Modelling A Discrete Event

Sometimes we wish to model a discrete event – e.g. whether a person obtains a degree, smokes or not, is employed or not, etc. In this case, we first model the individual age-specific probability of event occurring, and then – determine whether the event actually occurs by comparing the modelled probability with a random draw from a uniform distribution over a closed interval from zero to one. Literature researching discrete events most often reports estimates from a probabilistic regression, that represent either (i) average absolute change (percentage-point change) in the probability of the event occurring as a result of a unit change in the dependent variable; or (ii) odds ratio.

**Percentage Point Change.** Assume that we wish to model the effect of cognitive ability at age 18 (denoted $cog_{i,18}$) on whether an individual obtains university degree (denoted $edu_i$). Also, assume that it is known that a standard deviation change in the cognitive ability at age 18 increases the probability of obtaining a degree by $g$ percentage points. For example, Goodman et al. (2015) reports such estimates as average marginal effects from a probit regression model.[1]

[1] It should be noted that the average marginal effect is not always a good approximation of the true effect, as the actual individual marginal effect is not constant across individuals. So this method is a crude way of modelling the effect.

In a crude way, we can model the probability of obtaining a degree ($pr.edu_i$) as:

$$pr.edu_i = \overline{edu}[sex] + \frac{g}{100}\left(cog_{i,18} - \overline{cog}[18, sex]\right) + error_{i,19} \tag{4}$$

**Odds-Ratio.** When the effect estimates are obtained from a logistic probability regression model, they are often reported as odds ratios. For example, to assess the effect of depression on smoking, literature may report estimates of the following equation:

$$\ln\left(\frac{pr.smokes_{i,age}}{1 - pr.smokes_{i,age}}\right) = \alpha_{i,age} + \beta^{pr.smokes}_{depressed}\, depressed_{i,age} + error \tag{5}$$

where $depressed_{i,age}$ is an indicator of individual-depression; $\beta^{pr.smokes}_{depressed}$ is the natural logarithm of the reported odds ratio. Again, we can average equation (5), and as long as $ln\left(\frac{\overline{pr.smokes}[..]}{1-\overline{pr.smokes}[..]}\right)$ is a good approximation of $\frac{1}{N}\Sigma_i^N\{\ln\left(\frac{pr.smokes_{i,age}}{1-pr.smokes_{i,age}}\right)\}$, we can assume that $ln\left(\frac{\overline{pr.smokes}[..]}{1-\overline{pr.smokes}[..]}\right) = \overline{\alpha}[age, sex] + \beta^{pr.smokes}_{depressed}\overline{depressed}[..]$, where $\overline{pr.smokes}[..] \equiv \overline{pr.smokes}[age, sex]$, and $\overline{depressed}[..] \equiv \overline{depressed}[age, sex]$. We can express $\overline{\alpha}[age, sex]$ from this expression, and again assume that $\alpha_{age,i} = \overline{\alpha}[age, sex]$ and substitute $\overline{\alpha}[age, sex]$ in (5), then rearrange to get:

$$pr.smokes_{i,age} = \left(\frac{1}{\frac{\overline{pr.smokes}[..]}{1-\overline{pr.smokes}[..]}\exp\left(\beta^{pr.smokes}_{depressed}\left(depressed_{i,age} - \overline{depressed}[..]\right)\right)} + 1\right)^{-1} \tag{6}$$

#### A.2 Parameter Sources

Table A.1 explains the notation that we use to specify the modelling equations throughout the rest of the Appendix; Table A.2 summarises the target data; Table A.3 lists the literature sources of the parameter estimates used in parameterising the modelling equations; Table A.4 summarises what other variables these literature sources control for. We then provide full detailed specifications of the modelling equations to model each of the lifecourse outcomes, as well as full details on modelling taxes, cash benefits and costs associated with costly outcomes, in the next subsection.

**Table A.1: Notation**

| Notation | Explanation |
|---|---|
| <u>Simulated variables</u> | |
| *rec* | Recipient for the parent-training programme (indicator); |
| *cp* | Conduct problem measure; |
| *ip* | Impact of problems; |
| *cd* | Childhood conduct disorder (indicator); |
| *cog* | Cognitive skills; |
| *edu* | University degree (indicator); |
| *sm* | Smokes (indicator); |
| *dep* | Mental illness (indicator); |
| *chd* | Coronary heart disease (CHD) (indicator); |
| *dead* | Dead (indicator); |
| *pris* | In prison (indicator); |
| *care* | In residential care (indicator); |
| *empl* | Employed (indicator); |
| *earn* | Annual earnings, £; |
| *wealth* | Lifetime accumulated wealth; |
| *cons* | Annual consumption level, £; |
| *pov* | In poverty (indicator); |
| *tax* | Annual amount of taxes paid; |
| *benef* | Annual amount of benefits received; |
| *sav* | Savings rate; |
| *min.cons* | Minimum consumption level, which government subsidises if it cannot be sustained by an individual; |
| *sex* | Male (indicator); |
| *sep* | Socio-economic position (quintile group); |
| $sdq.cp.MCSj$ | SDQ conduct problem score reported in MCS sweep $j$ ($j = 2, 3, 4, 5, 6$); |
| $sdq.ip.MCSj$ | SDQ impact score reported in MCS sweep $j$; |
| $cog.MCSj$ | Extracted factor using principal component analysis based on cognitive skills tests reported in MCS sweep $j$, standardised with a mean of 1.00 and standard deviation of 0.15 following Jones and Schoon (2008); |
| <u>Other notation</u> | |
| prefix *pr.* | Probability, i.e. *pr.smokes* denotes probability of smoking; |
| line over variable (—) | Mean calculated from a target dataset, i.e. $\overline{smokes}[age, sex]$ is proportion of people smoking in a particular age and sex group; |
| prefix *trend.* | Modelled time trend, i.e. the mean increase in variable over time, estimated from a target dataset, i.e. during working years expected earnings increase as people get past their youth, as they gain work experience, climb the career ladder, etc.; |
| prefix *sd.* | Modelled variation in some variable, i.e. standard deviation in the variable, estimated from a target dataset; |
| $\beta_x^y$ | Parameter representing the effect of some outcome $x$ on some outcome $y$, i.e.$\beta_{sm}^{pr.chd}$ denotes the effect of smoking on CHD risk. Depending on the context, we use it to represent coefficients from a linear regression, odds-ratios, etc. See full list of parameters, and their sources in Table A.3; |
| $SMR_x$ | Standard mortality ratio given condition $x$, i.e. the probability of dying from condition $x$ divided by the probability of dying in the general population. |

Note: MCS – Millennium Cohort Study.

**Table A.2: Target Data**

| Parameter | Description | Source |
|---|---|---|
| $\overline{dead}[age, sex, sep]$ | Mortality rates (by age, sex and the English IMD quintile group); | ONS, 2011; |
| $\overline{dep}[age, sex, sep]$ | Ages 5-18: proportion of children with any emotional disorder (by age, sex, and IMD quintile group); age 18+: depression diagnosed by a doctor and present or being treated within the past 12 months in England (by age, sex, and English IMD quintile group); | For age 5-18: Mental Health of Children and Young People Great Britain, 2004; age 18+: Health Survey for England, 2014; |
| $\overline{chd}[age, sex, sep]$ | Proportion of people with CHD in England (by age, sex, and English IMD quintile group); | Health Survey for England, 2006; |
| $\overline{earn}[age, sex]$ | Mean full time annual gross pay in UK (by age and sex); | Annual Survey of Hours and Earnings, ONS, 2015; |
| $\overline{empl}[age, sex]$ | Seasonally adjusted employment rate, expressed as a proportion of the economically active population (by age and sex); | Labour Force Survey, ONS, 2018; |
| $\overline{sdq.cond}[age, sex]$ | Mean SDQ conduct problem score (by age and sex); | MCS, 2000-2014; |
| $\overline{cog}[age, sex]$ | Mean cognitive measure (by age and sex); | MCS, 2000-2014; |
| $\overline{edu}[19]$ | Higher Education Initial Participation Rate in 2015/2016 (estimate of the likelihood of a person participating in Higher Education by age 30, based on current participation rates, adjusted by the probability of dropping out); | Department for Education, 2016; |
| $\overline{sm}[14, sex]$ | Proportion of 14-year-old children smoking (by sex); | MCS, 2014; |
| $\overline{sm}[19, sex]$ | Proportion of daily smokers in England (by age, sex and English IMD quintile group) in England; | Health Survey for England, 2006; |
| $\overline{pov}[sex]$ | Proportion of households below 60% median income by sex in UK; | Family Resources Survey, Department for Work & Pensions, 2016/2017; |
| $\overline{cd}[4, sex]$ | Proportion of 4-year-old children with conduct disorder (by age, sex); | Mental Health of Children and Young People Great Britain, 2004; |
| $\overline{pris}[age, sex]$ | Average proportion of people in prison (by age and sex) in England and Wales over 31 March 2017 - 31 March 2018 (calculated using population estimates in mid-2017); | Offender Management Statistics, Ministry of Justice, 2017-2018; Population Estimates for UK, England and Wales, Scotland and Northern Ireland Mid-2017, ONS; |
| $\overline{care}[70, sex]$ | Proportion of people aged 65+ in resident care homes (by sex) in England and Wales, 2011; | "Changes in the older resident care home population between 2001 and 2011" 2014, ONS. |

Note: MCS – Millennium Cohort Study, ONS – Office for National Statistics, IMD – Index of Multiple Deprivation. Our notation uses an overline to denote averages from a target dataset.

**Table A.3: Parameters**

| Parameter | Value | Source | Notes |
|---|---|---|---|
| $SMR_{dep}$ | 3.21 among 15-44 year olds, 1.75 – 45-64 year olds and 1.18 for 65+ | Chang et al. (2010) | Age standardised mortality ratios in south-east London 2007-2009, for people with depressive episode against the general population of England and Wales in 2008; |
| $\beta^{pr.dead}_{chd}$ | See Table A.5 | Health survey for England (2006); the 20th Century Mortality Files, ONS; Mid-year population estimates for England and Wales, ONS | Estimated probability of dying from CHD among those who have a CHD, in England and Wales, 2008 using CHD prevalence rates of 2006; |
| $\beta^{earn}_{sdq.cond}$ | $\ln 1.004\,/SD_{soc}$ | Goodman et al. (2015) | 0.4% increase in gross wage with standard deviation increase in externalising subscale (conduct+peer); $SD_{soc}$ – standard deviation of SDQ conduct problem score in the relevant age-sex subgroup of our simulation. |
| $\beta^{earn}_{cog}$ | $\ln 1.072\,/SD_{cog}$ | Goodman et al. (2015) | 7.2% increase in gross wage with standard deviation increase in IQ score; $SD_{cog}$ – standard deviation of cognitive skills in the relevant age-sex subgroup of our simulation. |
| $\beta^{earn}_{edu}$ | $\ln 1.17$ if male; $\ln 1.37$ if female | Blundell et al. (2000) | 17% increase in hourly wage from having undergraduate degree for males, 37% for females; |
| $\beta^{pr.edu}_{cog}$ | $0.12/SD_{cog(19)}$ | Goodman et al. (2015) | standard deviation increase in cognitive ability associated with 12% point increase in prob. obtaining a degree; |
| $\beta^{pr.edu}_{sdq.cond}$ | $0.02/SD_{soc(19)}$ | Goodman et al. (2015) | standard deviation decrease in Rutter externalising score associated with 2.2% point increase in prob. obtaining a degree; |
| $\beta^{pr.edu}_{dep}$ | -0.04 | Goodman et al. (2015); Fletcher (2010); Farahati, Marcotte and Wilcox-Gök (2003) | Goodman et al. (2015) Fletcher (2010) find no statistically significant effect; but Fletcher (2010) finds that being depressed increases the probability of dropping out of high school by around 2.4% points, and decreases the probability of college enrolment by 2.7–7.2 percentage points. Farahati, Marcotte and Wilcox-Gök (2003) find that parent's depression increases child's probability of dropout by over 3% points for females. In the light of these findings, the current model specification sets the parameter at 4% points; |
| $\beta^{pr.sm}_{teen.sm}$ | $\ln 3.38$ if male; $\ln 3.68$ if female | Jefferis et al. (2003) | Estimates obtained using logistic regression; |
| $\beta^{pr.sm}_{pov}$ | $\ln 1.91$ if male; $\ln 1.81$ if female | Jefferis et al. (2003) | Estimates obtained using logistic regression; |
| $\beta^{pr.sm}_{edu}$ | $\ln 3.32$ if male; $\ln 3.26$ if female | Jefferis et al. (2003) | Estimates obtained using logistic regression; |

*Table continues on the next page.*

**Table A2: Parameters (*Continued*)**

| Parameter | Value | Source | Notes |
|---|---|---|---|
| $\beta_{dep}^{pr.sm}$ | $\ln 2.7$ | Lasser et al. (2000) | Estimates obtained using logistic regression; |
| $\beta_{pris}^{pr.sm}$ | 0.07 if male and 0.06 if female | Singleton, Farrell and Meltzer (2003) | Calculated using the prevalence rates in a population before and after imprisonment, does not take into account the contribution of this increase because of mental illness, poverty and potentially other variables; |
| $\beta_{cd}^{pr.dep}$ | $\ln 3.63$ | Luby et al. (2014) | Including the effect that occurs via non-supportive parenting (see discussion below); estimated using logistic regression; |
| $\beta_{unemploy}^{pr.dep}$ | $\ln 2.05$ if male; $\ln 1.72$ if female | Thomas, Benzeval and Stansfeld (2005) | Estimated using logistic regression; the effect on psychological problems measured by general health questionnaire; |
| $\beta_{employ}^{pr.dep}$ | $\ln 0.87$ if male; $\ln 0.79$ if female | Thomas, Benzeval and Stansfeld (2005) | Estimated using logistic regression; the effect on psychological problems measured by general health questionnaire; |
| $\beta_{pov}^{pr.dep}$ | $\ln 1.24$ | Weich and Lewis (1998) | Estimated using logistic regression; the effect on psychological problems measured by general health questionnaire; |
| $\beta_{pov}^{pr.chd}$ | $\ln 1.49$ if male; $\ln 1.18$ if female | Marmot et al. (1997) | Calculated using logistic regression controlling for age and CHD risk factors (incl. smoking), social support and job control. Using the parameters depends on assuming poverty correlates with low employment grade; |
| $\beta_{sm}^{pr.chd}$ | $\ln 2$ | Bazzano et al. (2003), Critchley and Capewell (2003) | Based on estimates of odds ratios reported in the cited sources (see discussion below); |
| $\beta_{cd}^{pr.pris}$ | 0.18 | Fergusson, John Horwood and Ridder (2005) | Estimated using rates of arrests/convictions among people with different levels of conduct problems; |
| $\beta_{dep}^{pr.prison}$ | 0.015 | Anderson, Cesur and Tekin (2015) | |
| $\beta_{dep}^{pr.care}$ | 0.18 | McDougall et al. (2007); Stewart et al. (2014) | Calculated using depression prevalence rates; |
| $\beta_{sdq.cond}^{pr.employ}$ | $0.016\,\beta_1/SD_{soc}$ | Goodman et al. (2015) | Standard deviation increase in externalising subscale of SDQ raises probability being employed by 1.6%; $SD_{soc}$ – standard deviation of SDQ conduct problem score in the relevant age-sex subgroup of our simulation; |
| $\beta_{cog}^{pr.employ}$ | $0.021\,\beta_1/SD_{cog}$ | Goodman et al. (2015) | Standard deviation increase in IQ test score raises probability being employed by 2.1%; $SD_{cog}$ – standard deviation of the cognitive skills measure in the relevant age-sex subgroup of our simulation. |

#### Table A.4: Modelled Variables and Controls

| Y | Effect parameter | Method | Parameter reported | Explanatory variables (X) in the modelling equation | | | | | | | | | | | | | | |
|---|---|---|---|---|---|---|---|---|---|---|---|---|---|---|---|---|---|---|
| | | | | Cond. prob. | CD | Cog. skills | Education | Smoking | Teen. smoking | Depression | CHD | Employment | Prison | Res. care | Poverty | Income | Age | Sex |
| | | | | SOCIAL | | | | | | | | | | | | | | |
| Education | Cond. prob. | Probit; | AME; | ✓ | | ✓ | | | | × | | | | | (✓) | (✓) | | (✓) |
| Education | Cog. skills | Probit; | AME; | ✓ | | ✓ | | | | × | | | | | (✓) | (✓) | | (✓) |
| Education | Depression | Probit; | AME; | × | | ✓ | | | | ✓ | | | | | (✓) | (✓) | (✓) | (✓) |
| Unemployment | Cond. prob. | Probit; | AME; | ✓ | | ✓ | | | | × | | | | | (✓) | (✓) | | (✓) |
| Unemployment | Cog. skills | Probit; | AME; | ✓ | | ✓ | | | | × | | | | | (✓) | (✓) | | (✓) |
| Prison | CD | Compare prevalence rates across subgroups, test the significance of relationships using logit; | Average rates of being arrested/convicted among the different subgroups; | (✓) | ✓ | (✓) | | | | × | | | | | (✓) | | | (✓) |
| Prison | Depression | OLS (robustness checks with probit and logit yield similar results); | Regression coefficient; | | ✓(control for drug, alcohol and marijuana use, ADHD, bad temper and anxiety during adolescence) | | (✓) | | (✓) | ✓ | | (✓) | | | (✓) | (✓) | (✓) | (✓) |
| Res. care | Depression | Compare prevalence rates across subgroups; | Age and sex adjusted difference between subgroups; | | | | | | | ✓ | | | | | | | (✓) | (✓) |

*Table continues on the next page.*

**Table A4: Modelled Variables and Controls (*Continued*)**

| Y | Effect parameter | Method | Parameter reported | Cond. prob. | CD | Cog. skills | Education | Smoking | Teen. smoking | Depression | CHD | Employment | Prison | Res. care | Poverty | Income | Age | Sex |
|---|---|---|---|---|---|---|---|---|---|---|---|---|---|---|---|---|---|---|
| | | | | | | | HEALTH | | | | | | | | | | | |
| Smokes | Education | Logit; | OR; | | | | ✓ | | ✓ | × | | | × | | × | | | (✓) |
| | Teen. smoking | Logit; | OR; | | | | × | | ✓ | × | | | × | | ✓(manual social class) | | | (✓) |
| | Poverty | Logit; | OR; | | | | × | | ✓ | × | | | × | | ✓ | | | (✓) |
| | Depression | Logit; | OR; | | | | × | | × | ✓ | | | × | | × | | (✓) | (✓) |
| | Prison | Comparison of smoking status pre and post imprisonment; | Increase in smoking rate post imprisonment; | | | | × | | × | × | | | ✓ | | × | | | (✓) |
| Depressed | CD | Logit; | OR; | | ✓ | | | | | | | | | | ✓(family income-to-needs ratio) | | (✓) | |
| | Unemployment | Logit; | OR; | | × | | | | | ✓(prior mental illness) | | ✓ | | | × | (✓) | (✓) | |
| | Poverty | Logit; | OR; | | × | | (✓) | | | | | ✓ | | | ✓ | (✓) | (✓) | (✓) |
| CHD | Smoking | Logit; | OR; | | | | | ✓ | | | | | | | × | | (✓) | (✓) |
| | Poverty | Logit; | OR; | | | | | ✓ | | | | | | | ✓(low employment grade) | | (✓) | (✓) |
| Mortality | Depression | Estimation of standardised mortality ratios; | Age standardised mortality ratio; | | | | | | | ✓ | × | | | | | | (✓) | (✓) |
| | CHD | Estimation of dying probability from CHD; | Probability of dying from CHD; | | | | | | | × | ✓ | | | | | | (✓) | (✓) |
| | | | | | | | ECONOMIC | | | | | | | | | | | |
| Earnings | Cond. prob. | Probit; | AME; | ✓ | | ✓ | | | | × | | | | | (✓) | (✓) | | (✓) |
| | Cog. skills | Probit; | AME; | ✓ | | ✓ | | | | × | | | | | (✓) | (✓) | | (✓) |
| | Education | Regression based linear matching; | Regression coefficient; | × | | | ✓ | × | | | | | | | ✓ | ✓ | | (✓) |

Note: ✓– variable X is included in the modelling equation for Y, as well as was controlled for in the literature; (✓) – variable X is not included in the modelling equation for Y, but indirectly influences Y through the other LifeSim equations, as well as was controlled for in the literature; ×– variable X is included in the modelling equation for Y, but was not controlled for in the literature; AME – average marginal effects; OLS – ordinary least squares. Other abbreviations: AME – average marginal effects, OR – odds ratio, cond. prob. – conduct problems, CD- conduct disorder, teen. smoking – teenage smoking, CHD – coronary heart disease, res. care – residential care.

#### A.3 Specification

We present the full specification of the modelling equations which follows the structure outlines in Table 3 in the main text. This material should be used together with Table A.1, which clarifies the notation, as well as Table A.2 in the main text, which specifies the target data sources, and Table A.3 and Table A.4, which specify the parameters, and details about their sources.

##### A.3.1 Skills Outcomes

**Conduct Problems.** Modelled using SDQ conduct problems scale data from the MCS.

$$\begin{cases} cp_{i,age} = sdq.cond.MCS2_i & \text{if } age \leq 4; \\ cp_{i,age} = sdq.cond.MCS3_i & \text{if } age \in [5,6]; \\ cp_{i,age} = sdq.cond.MCS4_i & \text{if } age \in [7,10]; \\ cp_{i,age} = sdq.cond.MCS5_i & \text{if } age \in [11,13]; \\ cp_{i,age} = sdq.cond.MCS6_i & \text{if } age \in [14,18]; \\ cp_{i,age} = n/a & \text{if } age \geq 19. \end{cases} \tag{7}$$

**Impact of Problems.** Modelled using SDQ impact supplement data from the MCS.

$$\begin{cases} ip_{i,age} = sdq.ip.MCS2_i & \text{if } age \leq 4; \\ ip_{i,age} = sdq.ip.MCS3_i & \text{if } age \in [5,6]; \\ ip_{i,age} = sdq.ip.MCS4_i & \text{if } age \in [7,18]; \\ ip_{i,age} = n/a & \text{if } age \geq 19. \end{cases} \tag{8}$$

**Cognitive Skills.** Modelled using principal component analysis to extract a common factor from the various cognitive skills measures disseminated by the MCS, following Jones and Schoon

(2008) standardised with a mean of 1.00 and standard deviation of 0.15.

$$\begin{cases} cog_{i,age} = cog.MCS2_i & \text{if } age \leq 4; \\ cog_{i,age} = cog.MCS3_i & \text{if } age \in [5,6]; \\ cog_{i,age} = cog.MCS4_i & \text{if } age \in [7,10]; \\ cog_{i,age} = cog.MCS5_i & \text{if } age \in [11,13]; \\ cog_{i,age} = cog.MCS6_i & \text{if } age \in [14,18]; \\ cog_{i,age} = n/a & \text{if } age \geq 19. \end{cases} \tag{9}$$

##### A.3.2 Social Outcomes

**Childhood Conduct Disorder.** Modelled using the predictive algorithm by Goodman et al. (2003); Goodman, Renfrew and Mullick (2000).

$$\begin{cases} pr.cd_{i,age} = 0.61 & \text{if } age \in [5,18] \ \& \ cp_{i,age} \geq 5 \ \& \ ip_{i,age} \geq 2 \\ pr.cd_{i,age} = 0.31 & \text{if } age \in [5,18] \ \& \ cp_{i,age} \geq 4; \\ pr.cd_{i,age} = 0.06 & \text{if } age \in [5,18] \ \& \ cp_{i,age} < 4; \\ pr.cd_{i,age} = n/a & \text{if } age \in [0,4] \text{ or } age > 18. \end{cases} \tag{10}$$

**Education (University Degree).** We model the probability of obtaining a university degree at age 19:[2]

$$\begin{cases} pr.edu_{i,age} = \max[0, \min[1, \overline{edu}[.] + \beta_{cog}^{pr.edu}\,(cog_{i,age-1} - \overline{cog}[.]) + \\ \quad +\beta_{cp}^{pr.edu}\,(10 - cp_{i,age-1} + \overline{cp}[.]) + \\ \quad +\beta_{dep}^{pr.edu}\,\big(dep_{i,age-1} - \overline{dep}[.]\big)\ ]\,] \qquad \text{if } age = 19; \\ pr.edu_{i,age} = n/a \qquad \text{if } age \neq 19. \end{cases} \tag{11}$$

where $\overline{edu}[..] \equiv \overline{edu}[age_i, sex_i]$, $\overline{cog}[.] \equiv \overline{cog}[age_i - 1, sex_i]$,$\overline{cp}[.] \equiv \overline{cp}[age_i - 1, sex]$, $\overline{dep}[.] \equiv \overline{dep}[age_i - 1, sex]$

[2]We assume that whether an individual obtains a university degree is determined at age 19.

**Unemployment/Employment.** During 'working years' we model the individual probability of being employed; if individual is in prison, he/she is not employed by definition and this probability is zero.

$$\begin{cases} pr.empl_{i,age} = n/a \quad \text{if } age \in [0,18] \text{ or } age \geq 70; \\ pr.empl_{i,age} = 0 \quad \text{if } pris_{i,age} = 1; \\ pr.empl_{i,age} = \max[0, \quad \min[1, \quad \overline{empl}[.]+ \\ \quad +\beta_{cp}^{pr.employ} \ (cp_{i,age-1} - \overline{cp}[.]) + \\ \quad +\beta_{cog}^{pr.empl} \ (cog_{i,age-1} - \overline{cog}[.]) \quad ]] \qquad \text{if } age = 19; \\ pr.empl_{i,age} = \max[0, \min[1, \ pr.empl_{i,age-1} + trend.\overline{empl}[.] \,]] \\ \quad \text{if } age \in [20, 69]. \end{cases} \tag{12}$$

where $\overline{empl}[.] \equiv \overline{empl}[age_i, sex_i]$ ,$\overline{cp}[.] \equiv \overline{cp}[age_i - 1, sex]$, $\overline{cog}[.] \equiv \overline{cog}[age_i - 1, sex_i]$.

**Poverty.** We model poverty as an indicator when individual consumption level falls below the absolute poverty line, 60% median equivalised household income in the UK in year 2011, which we set at £14,637 (Office for National Statistics).

**Prison.** During 'working years', individuals can go to prison, so we model the probability of being in prison. Imprisoned individuals are assumed to be unemployed and do not receive any salary; they are assumed to consume at a level equivalent to the state-subsidised minimum, which is subsidised by their own wealth (if sufficiently wealthy) or the state.

$$\begin{cases} pr.pris_{i,age} = n/a \qquad \text{if } age \in [0,18] \text{ or } age \geq 70; \\ pr.pris_{i,age} = \max[0, \min[1, \overline{pris}[.]+ \\ \quad +\beta_{cd}^{pr.pris} \left(cd_{i,age-1} - \overline{cd}[.]\right) + \beta_{dep}^{pr.pris} \left(dep_{i,age-1} - \overline{dep}[.]\right)]] \\ \quad \text{if } age = 19; \\ pr.pris_{i,age} = \max[0, \min[1, \\ \quad pr.pris_{i,age-1} + \beta_{dep}^{pr.pris} \triangle dep_{i,age-1}]] \qquad \text{if } age \in [20, 69]. \end{cases} \tag{13}$$

where $\overline{pris}[.] \equiv \overline{pris}[age_i, sex_i]$, $\overline{cd}[.] \equiv \overline{cd}[age_i - 1, sex_i]$ and $\overline{dep}[.] \equiv \overline{dep}[age_i - 1, sex_i]$.

**Residential Care.** During 'retirement', individuals can live in residential care home, so we model the probability of living in a care home. We assume that individuals cover their care home cost (denoted *care.cost*, see Table A.6), if they have sufficient resources to do so; otherwise, the state subsidises their care home cost.

$$\begin{cases} pr.care_{i,age} = n/a \qquad \text{if } age \leq 69; \\ pr.care_{i,age} = \max[0, \min[1, \overline{care}[.]+ \\ \qquad +\beta_{dep}^{pr.care}\left(dep_{i,age} - \overline{dep}[.]\right)]] \qquad \text{if } age = 70; \\ pr.care_{i,age} = \max[0, \min[1, pr.care_{i,age-1} + \beta_{dep}^{pr.care} \triangle dep_{i,age}]] \\ \qquad \text{if } age > 70. \end{cases} \tag{14}$$

where $\overline{care}[.] \equiv \overline{care}[age_i, sex_i]$ and $\overline{dep}[.] \equiv \overline{dep}[age_i, sex_i]$.

##### A.3.3 Health Outcomes

**Smoking.**

$$\begin{cases} pr.sm_{i,age} = n/a \quad \text{if } age \in [0, 18]; \\ pr.sm_{i,age} = \max[0, \min[1, \left(\left(\frac{\overline{sm}[.]}{(1-\overline{sm}[.])}\exp\Phi\right)^{-1} + 1\right)^{-1} + \\ \qquad +\beta_{pris}^{pr.sm}\left(pris_{i,age} - \overline{pris}[.]\right) \quad ]\ ] \qquad \text{if } age = 19\ ; \\ pr.sm_{i,age} = \max[0, \min[1, \\ \qquad \left(\frac{1}{\frac{pr.sm_{i,age-1}}{(1-pr.sm_{i,age-1})}\exp\left(\beta_{pov}^{pr.sm}\triangle pov_{i,age}+\beta_{dep}^{pr.sm}\triangle dep_{i,age}\right)} + 1\right)^{-1} + \\ \qquad +\beta_{pris}^{pr.sm}\triangle pris_{i,age} + trend.\overline{sm}[.] \quad ]\ ] \qquad \text{if } age \in [19, 69]; \\ pr.sm_{i,age} = \max[0, \min[1, \\ \qquad \left(\frac{1}{\frac{pr.sm_{i,age-1}}{(1-pr.sm_{i,age-1})}\exp\left(\beta_{pov}^{pr.sm}\triangle pov_{i,age}+\beta_{dep}^{pr.sm}\triangle dep_{i,age}\right)} + 1\right)^{-1} + \\ +trend.\overline{sm}[.]]] \\ \qquad \text{if } age \geq 70. \end{cases} \tag{15}$$

where $\Phi = \beta_{teen.sm}^{pr.sm}\left(sm_{i,14} - \overline{sm}[14, sex]\right) + \beta_{pov}^{pr.sm}\left(pov_{i,age-1} - \overline{pov}[.]\right) +$
$+ \beta_{edu}^{pr.sm}\left(edu_{i,age} - \overline{edu}[.]\right) + \beta_{dep}^{pr.sm}\left(dep_{i,age-1} - \overline{dep}[.]\right)$, and
$\overline{sm}[.] \equiv \overline{sm}[age, sex]$, $\overline{pris}[.] \equiv \overline{pris}[age, sex]$, $\overline{pov}[.] \equiv \overline{pov}[age-1, sex]$, $\overline{edu}[.] \equiv \overline{edu}[age, sex]$,

$\overline{dep}[.] \equiv \overline{dep}[age-1, sex]$

**Depression.**

$$\begin{cases} pr.dep_{i,age} = n/a \quad \text{if } age \leq 4; \\ pr.dep_{i,age} = \max[0, \min[1, \\ \quad \left( \frac{1}{\frac{\overline{dep}[age,sex]}{1-\overline{dep}[age,sex]} \exp\left(\beta_{cd}^{pr.dep}\left(cd_{i,age-1} - \overline{cd}[age-1,sex]\right)\right)} + 1 \right)^{-1} ]] \\ \quad \text{if } age = 5; \\ pr.dep_{i,age} = \max[0, \min[1, \\ \quad \left( \frac{1}{\frac{pr.dep_{i,age-1}}{1-pr.dep_{i,age-1}} \beta_{cd}^{pr.dep} \exp\left(\beta_{cd}^{pr.dep} \triangle cd_{i,age-1}\right)} + 1 \right)^{-1} + trend.\overline{dep}[age, sex]]] \\ \quad \text{if } age \in [6, 18]; \\ pr.dep_{i,age} = \max[0, \min[1, \\ \quad \left( \frac{1}{\frac{pr.dep_{i,age-1}}{1-pr.dep_{i,age-1}} \exp\left(-\beta_{unempl}^{pr.dep} \triangle empl_{i,age-1} + \beta_{pov}^{pr.dep} \triangle pov_{i,age-1}\right)} + 1 \right)^{-1} + \\ +trend.\overline{dep}[age, sex]]] \\ \quad \text{if } age \in [19, 69] \\ pr.dep_{i,age} = pr.dep_{i,age-1} \quad \text{if } age \geq 70. \end{cases} \tag{16}$$

**Coronary Heart Disease.**

$$\begin{cases} pr.chd_{i,age} = n/a \quad \text{if } age \in [0, 18]; \\ pr.chd_{i,age} = \max[0, \quad \min[1, \\ \quad \left( \frac{1}{\frac{\overline{chd}[.]}{1-\overline{chd}[.]} \exp\left(\beta_{sm}^{pr.chd}\left(sm_{i,age-1} - \overline{sm}[.]\right) + \beta_{pov}^{pr.chd}\left(pov_{i,age-1} - \overline{pov}[.]\right)\right)} + 1 \right)^{-1} ]] \\ \quad \text{if } age = 19; \\ pr.chd_{i,age} = \max[0, \quad \min[1, \\ \quad \left( \frac{1}{\frac{pr.chd_{i,age-1}}{1-pr.chd_{i,age-1}} \exp\left(\beta_{sm}^{pr.chd} \triangle sm_{i,age} + \beta_{pov}^{pr.chd} \triangle pov_{i,age}\right)} + 1 \right)^{-1} ]]+ \\ \quad +trend.\overline{chd}[.] \qquad \text{if } age \geq 20. \end{cases} \tag{17}$$

where $\overline{chd}[.] \equiv \overline{chd}[age_i, sex_i]$ ,$\overline{sm}[.] \equiv \overline{sm}[age_i - 1, sex]$, $\overline{pov}[.] \equiv \overline{pov}[age_i - 1, sex_i]$

**Mortality.**

$$\begin{cases} pr.dead_{i,age} = \overline{dead}[.] \quad \text{if } age \in [0,4]; \\ pr.dead_{i,age} = \max[0, \min[1, \overline{dead}[.]\,(1 + (smr_{dep} - 1)dep_{i,age})]] \\ \quad \text{if } age \in [5,18]; \\ pr.dead_{i,age} = \max[0, \min[1, \overline{dead}[.]\,(1 + (smr_{dep} - 1)dep_{i,age}) + \\ \quad + \beta_{chd}^{pr.dead}\left(chd_{i,age} - \overline{chd}[.]\right) \quad ]] \qquad \text{if } age > 18. \end{cases} \tag{18}$$

where $\overline{dead}[.] \equiv \overline{dead}[age_i, sex_i, sep_i]$ and $\overline{chd}[.] \equiv \overline{chd}[age_i, sex_i, sep_i]$.

**Table A.5: Mortality from Coronary Heart Disease**

| | Sex | Age band | | | | | | |
|---|---|---|---|---|---|---|---|---|
| | | 16-24 | 25-34 | 35-44 | 45-54 | 55-64 | 65-74 | 75+ |
| Mortality, | male | 0.19 | 1.24 | 2.81 | 1.83 | 1.61 | 2.07 | 5.31 |
| % | female | 0.06 | 0.49 | 1.26 | 1.13 | 1.43 | 1.95 | 8.82 |

Note: Estimated mortality from coronary heart disease (CHD) among people diagnosed with CHD. These estimates are used to model the parameter $\beta_{chd}^{pr.dead}$ in equation (18) and Table A.3.

##### A.3.4 Economic Outcomes

**Earnings from Employment.** We model the gross annual salary for people who are employed.

$$\begin{cases} earn_{i,age} = n/a \qquad \text{if } age \in [0,18] \text{ or } age \geq 70; \\ earn_{i,age} = 0 \qquad \text{if } age \in [19,70] \;\&\; empl_{i,age} = 0\;; \\ earn_{i,19} = \max[0, \\ \quad (\overline{earn}[.] + sd.\overline{earn}[.])\, \exp(\beta_{cp}^{earn}\,(10 - cp_{i,age-1} + \overline{cp}[.]) + \\ \quad + \beta_{cog}^{earn}\,(cog_{i,age-1} - \overline{cog}[.]) + \beta_{edu}^{earn}\,(edu_{i,age} - \overline{edu}[.])) \quad ] \\ \quad \text{if } age = 19 \;\&\; employ_{i,19} = 1; \\ earn_{i,age} = \max[0, \quad earn_{i,age-h} + trend.\overline{earn}[.] \quad ] \\ \quad \text{if } age \in [20,69] \;\&\; empl_{i,age} = 1. \end{cases} \tag{19}$$

where '$h$' is years since individual $i$ was last employed, or $age - 19$, if individual was never employed (in this case we use the value of individual's potential earnings at 19); $\overline{earn}[.] \equiv$

$\overline{earn}[age_i, sex_i]$, $\overline{cp}[.] \equiv \overline{cp}[age_i - 1, sex_i]$, $\overline{cog}[.] \equiv \overline{cog}[age_i - 1, sex_i]$ and $\overline{edu}[.] \equiv \overline{edu}[age_i, sex_i]$.

**Earnings from Interest (Interest).** See details in the section below on modelling "Wealth".

**Pension.** Individuals are assumed to receive equivalent to the new basic UK state pension, if they have been employed for at least 10 years.[3].

**Savings.** During 'working-years', some individuals save part of their annual net income (i.e. earnings from employment and interest, net of tax). It is assumed that individuals save as much as they can, given that (a) they can sustain at least the previous year's consumption level and (b) their total annual savings do not exceed 16% of their annual net income.[4] This means that individuals who experience a negative income shock, reduce their savings relative to previous year to sustain their consumption level; individuals who experience a positive income shock and can afford to consume at least the level of government subsidised minimum '*min.cons*'[5] – save a positive amount up to 16% net income; finally, individuals who experience a high positive income shock, and who could potentially afford saving above 16% while still sustaining previous year's consumption – consume anything excess of 16% of their net disposable income (instead of saving it).

$$\begin{cases} sav_{i,age} = 0 \qquad \text{if } age \in [0, 18] \text{ or } age \geq 70; \\ sav_{i,age} = \max[0, \min[\ \ 0.16 \times (earn_{i,age} + int_{i,age} - tax_{i,age}), \\ \qquad earn_{i,age} + int_{i,age} - tax_{i,age} - cons_{i,age-1}\ \ ]\ \ ] \\ \qquad \text{if } age \in [19, 69]. \end{cases} \tag{20}$$

**Family Wealth.** At the age 19, individuals are assumed to inherit wealth from parents. Wealth generates annual interest, i.e. $int_{i,age} = wealth_{i,age-1} \times (1 + r)$, where $r$ is the annual interest rate, which we set at 1%. During their 'working years' individuals accumulate additional wealth by saving, if they can afford to save. If individuals experience a negative income shock,

[3]See details on UK new State Pension at `https://www.gov.uk/new-state-pension`

[4]This threshold is chosen as a maximum, informing from historical UK households savings ratios reported by ONS: `https://www.ons.gov.uk/economy/grossdomesticproductgdp/timeseries/dgd8/ukea`.

[5]See the paragraph below about "Consumption" in Section A.3.5.

they use their wealth to subsidise their consumption.

$$\begin{cases} wealth_{i,age} = \max[0, par.wealth_{i,age-1} + int_{i,age} + \\ \quad + \min[0, par.inc_i - cons_{i,age}]] \qquad \text{if } age \leq 18; \\ wealth_{i,age} = \max[0, \quad wealth_{i,age-1} + \\ \quad + \min[sav_{i,age}, \quad earn_{i,age} + int_{i,age} - tax_{i,age} - cons_{i,age}]] \\ \quad \text{if } age \in [19, 69]; \\ wealth_{i,age} = \max[0, \quad wealth_{i,age-1} + int_{i,age} + pens_{i,age} - \\ \quad - cons_{i,age} - tax_{i,age} - care_{i,age} \times care.cost] \\ \quad \text{if } age \geq 70. \end{cases} \tag{21}$$

where $par.wealth$ – parental wealth and $par.inc$ – parental income, as given in the childhood dataset.

**Taxes.** Individuals pay annual taxes on their income, i.e. earnings from employment and interest, as well as pension. The individual tax rate is set according to the corresponding UK tax bracket.[6]

**Benefits.** Individuals receive benefits subsidised by the public budget ($benef_{i,age}$) to sustain the minimum consumption level of £10,000, whenever they cannot afford it from their own net income (parental income and interest during 'pre-school years' and 'school years', salary and interest during 'working years', and pension and interest during 'retirement') and wealth. During 'retirement', individuals also receive benefits when in care to cover the care home costs, when they do not have sufficient own resources to cover them.

##### A.3.5 Wellbeing Outcomes

**Consumption.** It is assumed that government subsidises consumption level of at least '$min.cons$' (the state-subsidised minimum), in the case when individual cannot afford it given their income or wealth. We set $min.cons = \pounds 10,000$.

Up to age 18, individuals are assumed to consume the level of their household equivalised income

[6]See UK income tax rates at `https://www.gov.uk/income-tax-rates`. We use the year 2018/19 rates, converted to year 2015/16 prices.

($par.inc$) as given in the childhood dataset, or the state-subsidised minimum.

During ‘working years’ individuals consume what is left of their income from employment and interest after tax and savings, or an amount equal to the state-subsidised minimum (this may be subsidised by state or own wealth, depending on whether individual has positive wealth). For more details, read about the savings equation above.

During ‘retirement’, individuals try to sustain their previous year’s consumption level if they can afford it given their resources (i.e. net income from interest, state pension, their wealth and minus residential care home cost, if in care); if individuals cannot afford sustaining previous year’s consumption level, then they consume the maximum amount that they can afford given their resources, or the state-subsidised minimum.

$$\begin{cases} cons_{i,age} = \max[min.cons, par.inc_i] \qquad \text{if } age \leq 18; \\ cons_{i,age} = \max[min.cons, earn_{i,age} + int_{i,age} - tax_{i,age} - \\ \quad -sav_{i,age}] \qquad \text{if } age \in [19, 69]; \\ cons_{i,age} = \max[min.cons, \min[cons_{i,age-1}, wealth_{i,age-1} + \\ \quad +int_{i,age} - tax_{i,age} + pens_{i,age} - care_{i,age} \times care.cost] \quad ] \\ \quad \text{if } age \geq 70. \end{cases} \tag{22}$$

**Health Quality.** Health quality depends on the two health outcomes that we model – mental illness (depression) and physical illness (CHD) – as well as the aggregate health quality in England. More specifically, $health_{i,age} = h(chd_{i,age}, dep_{i,age})$, where $h(.)$ is a function decreasing in negative health experiences, and with a maximum of 1 when individual is in full health and anchored at 0 when individual is dead or in a health state as bad as death. More specifically, we assume $h(..) = \min[1, \max[0, \overline{health}[age, sex, sep] - (d(chd) \times chd_{i,age} + d(dep) \times dep_{i,age})]]$, where $\overline{health}[age, sex, sep]$ is the average health quality in England by age, sex and English IMD quintile group (Love-Koh et al., 2015), $d(x)$ represents the excess reduced health quality from the health condition $x$ (we use data for health quality with affective disorders and coronary atherosclerosis from Sullivan et al. (2011)).

##### A.3.6 Public Costs and Revenues

We model the costs associated with different outcomes, as summarised in table A.6. We assume that the following outcomes incur costs to the public service: CHD, depression, other healthcare, conduct disorder, prison, residential care.

**Table A.6: Public Service Costs**

| Cost type | Components of the cost | Annual cost per person, £ | Source |
|---|---|---|---|
| Healthcare: Coronary heart disease[7] | Direct health care cost; | 840; | Liu et al. (2002); |
| | Informal care cost; | 1,173; | |
| Healthcare: Depression | Costs to the National Health Service, the Accident and Emergency department, other support services (average); | 5,260; | McCrone et al. (2008); |
| Other healthcare | Average English National Health Service healthcare spending in the financial year 2011/12 by age, sex and English neighbourhood deprivation quintile group; | see Asaria (2017); | Asaria (2017); |
| Conduct disorder | Cost to the National Health Service; | 1,243 (age 5-10), 113 (age 11+); | Edwards et al. (2007); Scott et al. (2001), cited by Bonin et al. (2011); |
| | Cost to the Social Services Department; | 175 (age 5-10), 70 (age 11+); | Edwards et al. (2007); Romeo, Knapp and Scott (2006), cited by Bonin et al. (2011); |
| | Cost to the Department for Education; | 985 (age 5-10), 1,3402 (age 11-16), 0 (age 17+); | Edwards et al. (2007); Scott et al. (2001), cited by Bonin et al. (2011); |
| | Cost to the voluntary Sector; | 26; | Edwards et al. (2007), cited by Bonin et al. (2011); |
| Prison | Unit annual costs of custody (per year); | 31,925; | |
| | Unit costs of police (per record crime); | 553; | Dubourg et al. (2005); |
| | Unit costs of courts (per court event); | 7,103; | |
| Residential care | Cost of residential home; | 29,934; | Curtis and Burns (2017). |

Note: We uprate all the costs to year 2015/16 prices.

#### Appendix B References for Data and Parameter Sources

**Anderson, D Mark, Resul Cesur, and Erdal Tekin.** 2015. "Youth Depression and Future Criminal Behavior." *Economic Inquiry*, 53(1): 294–317.

**Asaria, Miqdad.** 2017. "Health Care Costs in the English NHS: Reference Tables for Average Annual NHS Spend by Age, Sex and Deprivation Group." Centre for Health Economics at the University of York Research Paper 147.

**Bazzano, Lydia A, Jiang He, Paul Muntner, Suma Vupputuri, and Paul K Whelton.** 2003. "Relationship Between Cigarette Smoking and Novel Risk Factors for Cardiovascular Disease in the United States." *Annals of Internal Medicine*, 138(11): 891–897.

**Blundell, Richard, Lorraine Dearden, Alissa Goodman, and Howard Reed.** 2000. "The Returns to Higher Education in Britain: Evidence From a British Cohort." *The Economic Journal*, 110(461): 82–99.

**Bonin, Eva-Maria, Madeleine Stevens, Jennifer Beecham, Sarah Byford, and Michael Parsonage.** 2011. "Costs and Longer-Term Savings of Parenting Programmes for the Prevention of Persistent Conduct Disorder: A Modelling Study." *BMC Public Health*, 11(1): 803.

**Chang, Chin-Kuo, Richard D Hayes, Matthew Broadbent, Andrea C Fernandes, William Lee, Matthew Hotopf, and Robert Stewart.** 2010. "All-Cause Mortality Among People With Serious Mental Illness (SMI), Substance Use Disorders, and Depressive Disorders in Southeast London: A Cohort Study." *BMC Psychiatry*, 10(1): 77.

**Critchley, Julia A, and Simon Capewell.** 2003. "Mortality Risk Reduction Associated With Smoking Cessation in Patients With Coronary Heart Disease: A Systematic Review." *Journal of the American Medical Association*, 290(1): 86–97.

**Curtis, Lesley A, and Amanda Burns.** 2017. "Unit Costs of Health and Social Care 2017." Canterbury, United Kingdom: Personal Social Services Research Unit, University of Kent. `https://www.pssru.ac.uk/project-pages/unit-costs/unit-costs-2017/`. Accessed on 2020-02-26.

**Dubourg, Richard, Joe Hamed, Jamie Thorns, et al.** 2005. "The economic and social costs of crime against individuals and households 2003/04." *Home Office Online Report*, 30(05).

**Edwards, Rhiannon T, Alan Céilleachair, Tracey Bywater, Dyfrig A Hughes, and Judy Hutchings.** 2007. "Parenting Programme for Parents of Children at Risk of Developing Conduct Cisorder: Cost Effectiveness Analysis." *British Medical Journal*, 334(7595): 682.

**Farahati, Farah, Dave E Marcotte, and Virginia Wilcox-Gök.** 2003. "The Effects of Parents' Psychiatric Disorders on Children's High School Dropout." *Economics of Education Review*, 22(2): 167–178.

**Fergusson, David M, L John Horwood, and Elizabeth M Ridder.** 2005. "Show Me the Child at Seven: The Consequences of Conduct Problems in Childhood for Psychosocial Functioning in Adulthood." *Journal of Child Psychology and Psychiatry*, 46(8): 837–849.

**Fletcher, Jason M.** 2010. "Adolescent Depression and Educational Attainment: Results Using Sibling Fixed Effects." *Health Economics*, 19(7): 855–871.

**Gardner, Frances, Patty Leijten, Joanna Mann, Sabine Landau, Victoria Harris, Jennifer Beecham, Eva-Maria Bonin, Judy Hutchings, and Stephen Scott.** 2017. "Could Scale-Up of Parenting Programmes Improve Child Disruptive Behaviour and Reduce Social Inequalities? Using Individual Participant Data Meta-Analysis to Establish for Whom Programmes Are Effective and Cost-Effective." *Public Health Research*, 5(10).

**Goodman, Alissa, Heather Joshi, Bilal Nasim, and Claire Tyler.** 2015. *Social and Emotional Skills in Childhood and Their Long-Term Effects on Adult Life.* London, United Kingdom: Institute of Education. `https://www.nuffieldfoundation.org/news/active-policy-required-avoid-covid-19-crisis-exacerbating-inequalities` Accessed on 2020-10-28.

**Goodman, Robert, D Renfrew, and M Mullick.** 2000. "Predicting Type of Psychiatric Disorder From Strengths and Difficulties Questionnaire (SDQ) Scores in Child Mental Health Clinics in London and Dhaka." *European Child & Adolescent Psychiatry*, 9(2): 129–134.

**Goodman, Robert, Tamsin Ford, Helen Simmons, Rebecca Gatward, and Howart Meltzer.** 2003. "Using the Strengths and Difficulties Questionnaire (SDQ) to Screen for

Child Psychiatric Disorders in a Community Sample." *International Review of Psychiatry*, 15(1-2): 166–172.

**Jefferis, Barbara, Hilary Graham, Orly Manor, and C Power.** 2003. "Cigarette Consumption and Socio-Economic Circumstances in Adolescence as Predictors of Adult Smoking." *Addiction*, 98(12): 1765–1772.

**Lasser, Karen, J Wesley Boyd, Steffie Woolhandler, David U Himmelstein, Danny McCormick, and David H Bor.** 2000. "Smoking and Mental Illness: A Population-Based Prevalence Study." *Journal of the American Medical Association*, 284(20): 2606–2610.

**Liu, Joseph L Y, Nikolaos Maniadakis, Andrew Gray, and Mike Rayner.** 2002. "The Economic Burden of Coronary Heart Disease in the UK." *Heart*, 88(6): 597–603.

**Love-Koh, James, Miqdad Asaria, Richard Cookson, and Susan Griffin.** 2015. "The Social Distribution of Health: Estimating Quality-Adjusted Life Expectancy in England." *Value in Health*, 18(5): 655–662.

**Luby, Joan L, Michael S Gaffrey, Rebecca Tillman, Laura M April, and Andy C Belden.** 2014. "Trajectories of Preschool Disorders to Full DSM Depression at School Age and Early Adolescence: Continuity of Preschool Depression." *American Journal of Psychiatry*, 171(7): 768–776.

**Marmot, Michael G, Hans Bosma, Harry Hemingway, Eric Brunner, and Stephen Stansfeld.** 1997. "Contribution of Job Control and Other Risk Factors to Social Variations in Coronary Heart Disease Incidence." *The Lancet*, 350(9073): 235–239.

**McCrone, Paul R, Sujith Dhanasiri, Anita Patel, Martin Knapp, and Simon Lawton-Smith.** 2008. *Paying the Price: The Cost of Mental Health Care in England to 2026*. London, United Kingdom: King's Fund.

**McDougall, Fiona A, Kari Kvaal, Fiona E Matthews, Eugene Paykel, Peter B Jones, Michael E Dewey, and Carol Brayne.** 2007. "Prevalence of Depression in Older People in England and Wales: The MRC CFA Study." *Psychological Medicine*, 37(12): 1787–1795.

**Romeo, Renee, Martin Knapp, and Stephen Scott.** 2006. "Economic Cost of Severe Antisocial Behaviour in Children-And Who Pays It." *The British Journal of Psychiatry*, 188(6): 547–553.

**Scott, Stephen, Martin Knapp, Juliet Henderson, and Barbara Maughan.** 2001. "Financial Cost of Social Exclusion: Follow up Study of Antisocial Children into Adulthood." *British Medical Journal*, 323(7306): 191.

**Singleton, Nicola, Michael Farrell, and Howard Meltzer.** 2003. "Substance Misuse among Prisoners in England and Wales." *International Review of Psychiatry*, 15(1-2): 150–152.

**Stewart, Robert, Matthew Hotopf, Michael Dewey, Clive Ballard, Jatinder Bisla, Maria Calem, Viola Fahmy, Jo Hockley, Julie Kinley, Hywel Pearce, et al.** 2014. "Current Prevalence of Dementia, Depression and Behavioural Problems in the Older Adult Care Home Sector: The South East London Care Home Survey." *Age and Ageing*, 43(4): 562–567.

**Sullivan, Patrick W, Julia F Slejko, Mark J Sculpher, and Vahram Ghushchyan.** 2011. "Catalogue of EQ-5D Scores for the United Kingdom." *Medical Decision Making*, 31(6): 800–804.

**Thomas, Claudia, Michaela Benzeval, and Stephen A Stansfeld.** 2005. "Employment Transitions and Mental Health: An Analysis from the British Household Panel Survey." *Journal of Epidemiology & Community Health*, 59(3): 243–249.

**Weich, Scott, and Glyn Lewis.** 1998. "Material Standard of Living, Social Class, and the Prevalence of the Common Mental Disorders in Great Britain." *Journal of Epidemiology & Community Health*, 52(1): 8–14.
